# Laboratory Validation and Clinical Implementation of Human Monkeypox Saliva-Based Tests

**DOI:** 10.1101/2022.08.08.22278498

**Authors:** Lao-Tzu Allan-Blitz, Kevin Carragher, Adam Sukhija-Cohen, Hong Li, Jeffrey D. Klausner

## Abstract

**Background:** Improved diagnostic capabilities and accessibility are essential for controlling the outbreak of Human Monkeypox.

**Methods:** We describe a saliva-based polymerase chain reaction (PCR) assay for Human Monkeypox, *in vitro* test performance, and clinical implementation of that assay at three testing sites in Los Angeles. Finally, using pre-specified search terms, we conducted a systematic rapid review of PubMed and Web of Science online databases of studies reporting the performance of oral pharyngeal or saliva-based tests for Human Monkeypox.

**Results:** Laboratory evaluation of the assay showed in silico inclusivity of 100% for 97 strains of Human Monkeypox, with an analytic sensitivity of 250 copies/mL, and 100% agreement compared to known positive and negative specimens. Clinical testing identified 22 cases of Human Monkeypox among 132 individuals (16.7%). Of those 22 cases, 16 (72.7%) reported symptoms, 4 (18.2%) without a rash at the time of testing. Our systematic rapid review identified 6 studies; 23 patients had tests performed on oropharyngeal specimens 100% agreed with the PCR test result of a lesion swab.

**Conclusion:** Saliva-based PCR tests are potential tools for outbreak control, and further evaluation of the performance of such tests is warranted.

## Introduction

With cases of Human Monkeypox reported from 47 countries, the World Health Organization recently declared the current spread the infection a global emergency (1). Historically, Human Monkeypox has been endemic in tropical rainforest regions of Central and West Africa, with short-lived outbreaks driven by transmission through animal-to-human and human-to-human exposures (2). However, the current outbreak is now spreading much more rapidly and pervasively than any previous outbreak, with a unique pattern similar to a sexually transmitted disease (3-5). Such transmission has contributed to the disproportional burden of disease among men who have sex with men (4).

The Centers for Disease Control and Prevention has stressed the need for timely diagnosis as a primary means for outbreak control, particularly in the absence of sufficient vaccine supplies (6). Polymerase chain reaction (PCR) lesion testing was thought to be necessary, and the United States Food and Drug Administration has discouraged all testing apart from lesion swabs (7). The U.S. Centers for Disease Control and Prevention has given similar guidance (8).

Prior studies, however, suggest that viral DNA may be detected in saliva and oropharyngeal specimens (2, 9, 10). One recent report noted 100% (n=12) patients with Human Monkeypox had positive saliva PCR tests (9). Similar findings were reported among a study of 7 individuals diagnosed with Human Monkeypox in 2018 and 2019 (10). In both studies, many individuals had positive oropharyngeal tests early in the disease course (9, 10). Thus, via early case detection, saliva testing may provide the opportunity to limit infectiousness. We therefore assessed the performance of a newly-developed saliva-based PCR assay. We further report real-world implementation of that assay in clinical settings.

## Methods

We describe a PCR assay of saliva for the diagnosis of Human Monkeypox developed using DNA targets of Human Monkeypox genomes (Supplement 1). Patient saliva samples were collected to half of the collection kit volume (Supplement 2) and processed per the laboratory protocol. After collection, transport and receipt in the laboratory, specimens were washed. DNA was then extracted using the KingFisher™ Flex Purification System (ThermoFisher Scientific, Waltham, MA, USA). DNA amplification and detection was done using TaqMan^®^ Real-Time PCR (ThermoFisher Scientific, Waltham, MA, USA).

We report the microbiological inclusivity of all strains with genome sequences available, from two different clades (the West African Clade and the Central African Clade). We further report the analytic specificity of the assay for Human Monkeypox compared to other members of the orthopoxvirus genus as well as other non-orthopoxvirus genera. Subsequently, using 20 replicates of a positive control within pooled negative saliva (oral saliva matrix) specimens, we report the limit of detection, defined as the lowest concentration providing a positive result for 100% of replicates. Using two known positive specimens as well as 20 known negative specimens, we report the *in vitro* agreement between those specimens and our assay. For measures of agreement, at least two different operators and instruments were used on three separate days at three different concentrations to assess reproducibility (see Supplement 3). Assay validation was done in accordance with the United States Clinical Laboratory Improvement Act guidelines (11).

We reviewed de-identified patient records among individuals presenting for Human Monkeypox testing at saliva collection sites. Advarra institutional review committee exempted the analysis of de-identified data from institutional review (Pro00065270).

Finally, we conducted a systematic rapid review of the literature on PubMed and Web of Science databases to assess the performance of saliva tests in comparison to PCR of lesion swabs. We used the following pre-defined search terms: “monkeypox” AND (“diagnosis” OR “diagnostic”) AND (“saliva” OR “sputum” OR “throat” OR “pharyn*”). We further evaluated the references of all articles identified and searched pre-print servers for forthcoming publications. We included articles that reported the results of any oropharyngeal or saliva PCR tests for Monkeypox in humans. We excluded review articles, studies among primates, and studies not in English. We then conducted a narrative review of the studies, reporting individual study-level summary data given the degree of heterogeneity within studies precluded a formal meta-analysis.

## Results

The PCR saliva assay had an *in silico* inclusivity of 100% for all (n=97) strains from the two different clades. The assay was specific to the orthopoxvirus genus, but not to Human Monkeypox, as the assay also detected cowpox and rabbitpox, but did not detect non-orthopoxvirus genera. The analytic sensitivity was 250 copies/mL, the lowest dilution for which all (n=20) replicates were positive (Table 1). Further, there was 100% agreement compared to known positive and negative specimens over three separate days, performed by two different operators.

**Table 1:** *In Vitro* Limit of Detection for Saliva-Based PCR for Human Monkeypox via Serial Dilutions.

| Copies/mL | Replicates | Cycle Threshold Value | Interpretation | % Viral Positivity |
| --- | --- | --- | --- | --- |
| 0 | 1 | Undetermined | Negative | 0.0% |
|  | 2 | Undetermined | Negative |  |
|  | 3 | Undetermined | Negative |  |
| 31.25 | 1 | Undetermined | Negative | 33.3% |
|  | 2 | 35.7 | Positive |  |
|  | 3 | Undetermined | Negative |  |
| 62.5 | 1 | 34.1 | Positive | 66.7% |
|  | 2 | 34.11 | Positive |  |
|  | 3 | Undetermined | Negative |  |
| 125.0 | 1 | 36.34 | Positive | 66.7% |
|  | 2 | Undetermined | Negative |  |
|  | 3 | 34.8 | Positive |  |
| 250.0 | 1 | 34.8 | Positive | 100% |
|  | 2 | 34.6 | Positive |  |
|  | 3 | 35.7 | Positive |  |
| 500.0 | 1 | 34.7 | Positive | 100% |
|  | 2 | 32.6 | Positive |  |
|  | 3 | 34.1 | Positive |  |
| 1000.0 | 1 | 33.0 | Positive | 100% |
|  | 2 | 32.6 | Positive |  |
|  | 3 | 33.4 | Positive |  |
| 2000.0 | 1 | 31.9 | Positive | 100% |
|  | 2 | 31.9 | Positive |  |
|  | 3 | 31.5 | Positive |  |
| Controls |  |  |  |  |
| Positive Control 1 | 1 | 17.8 | Positive | 100% |
| Positive Control 2 | 1 | 28.7 | Positive | 100% |
| Positive Control 3 | 1 | 17.5 | Positive | 100% |
| Negative Control 1 | 1 | Undetermined | Negative | 0.0% |
| Negative Control 2 | 1 | Undetermined | Negative | 0.0% |
| Negative Control 3 | 1 | Undetermined | Negative | 0.0% |
| Negative Extra Control 1 | 1 | Undetermined | Negative | 0.0% |
| Negative Extra Control 2 | 1 | 39.4 | Negative | 0.0% |
| Negative Extra Control 3 | 1 | Undetermined | Negative | 0.0% |

Clinical testing from three testing sites in Los Angeles identified 22 cases of Human Monkeypox among 132 individuals screened (16.7%). Of those 22 patients, 16 (72.7%) reported symptoms, and 4 (18.2%) did not have a rash at the time of testing, while one patient reported being asymptomatic (Table 2). We did not have data on reported symptom status for five patients.

**Table 2:** Characteristics of 22 Cases of Human Monkeypox Identified Via Saliva-Based Testing at Three Publicly Accessible Testing Sites in Los Angeles.

| Case ID | Test Result | Cycle Threshold Value | Gender | Symptoms Reported | Known Exposure | No. Days Between Exposure and Symptom Onset | No. Days Between Symptom Onset and Positive Saliva Test |
| --- | --- | --- | --- | --- | --- | --- | --- |
| 1 | Positive | 27 | Male | Lesions, Headache, Fever, Sore Throat | Yes | 16 days | 1 day |
| 2 | Positive | 24 | Male | No Symptoms | Yes | - | - |
| 3 | Positive | 18 | Male | Lesions, Painful Lesions and/or Blisters, Headache, Fever, Muscle Pain, Sore Throat, Hot and Cold Flashes | Yes | 5 days | 8 days |
| 4 | Positive | 22 | Male | Lesions, Headache, Fever, Muscle Pain, Sore Throat, Hot and Cold Flashes | No known Exposure | - | 8 days |
| 5 | Positive | 32 | Male | Lesions, Headache, Muscle Pain, Sore Throat, Hot and Cold Flashes | Suspected Exposure | - | 15 days |
| 6 | Positive | 15 | Male | Sore Throat, Hot and Cold Flashes | Suspected Exposure | - | 4 days |
| 7 | Positive | 23 | Male | Lesions, Painful Lesions and/or Blisters, Headache, Fever, Muscle Pain, Hot and Cold Flashes | No known Exposure | - | 3 days |
| 8 | Positive | 16 | Male | Lesions, Headache, Fever, Sore Throat, Hot and Cold Flashes | Yes | 8 days | 2 days |
| 9 | Positive | 28 | Male | External Lesions (Hands, Face, Arms, Legs, Genital), Internal Lesions (Oral, Rectal), Headache, Fever | Yes | 3 days | 9 days |
| 10 | Positive | 21 | Male | External Lesions (Hands, Face, Arms, Legs, Genital), Headache, Fever, Chills and Hot Flashes, Diarrhea | No known Exposure | - | 5 days |
| 11 | Positive | 31 | Male | Fever, Muscle pain | No known Exposure | - | 8 days |
| 12 | Positive | 25 | Male | Lesions | Yes | - | 10 days |
| 13 | Positive | 31 | Male | - | - | - | - |
| 14 | Positive | 23 | Male | - | - | - | - |
| 15 | Positive | 28 | Male | - | - | - | - |
| 16 | Positive | 17 | Male | Lesions, painful lesions and/or blisters, headache, fever, muscle pain, sore throat, hot and cold flashes | Yes | 3 days | 35 days |
| 17 | Positive | 17 | Male | Headache, fever, muscle pain, sore throat, hot and cold flashes | Suspected Exposure | 7 days | 13 days |
| 18 | Positive | 22 | Male | Lesions, fever, muscle pain | Suspected Exposure | 5 days | 4 days |
| 19 | Positive | 21 | Male | - | - | - | - |
| 20 | Positive | 16 | Male | - | - | - | - |
| 21 | Positive | 20 | Male | Headache, Muscle pain, Sore throat | Yes | 6 days | 8 days |
| 22 | Positive | 18 | Male | Lesions | - | - | - |

Our systematic rapid review identified 16 articles, of which 6 meet our inclusion criteria, however one study by *Thornhill et al*. did not report numeric values for total number of oral swabs PCR tests performed, and thus was excluded. One further article was identified through evaluating references of the identified articles. Among those 6 studies (all case series) there were 292 total patients, 24 of whom had tests performed on oropharyngeal or saliva specimens (Table 3). In all included studies, the results of oral fluid specimen tests were positive among 100% of patients with concomitant positive lesion swabs for human monkeypox infection.

**Table 3:**
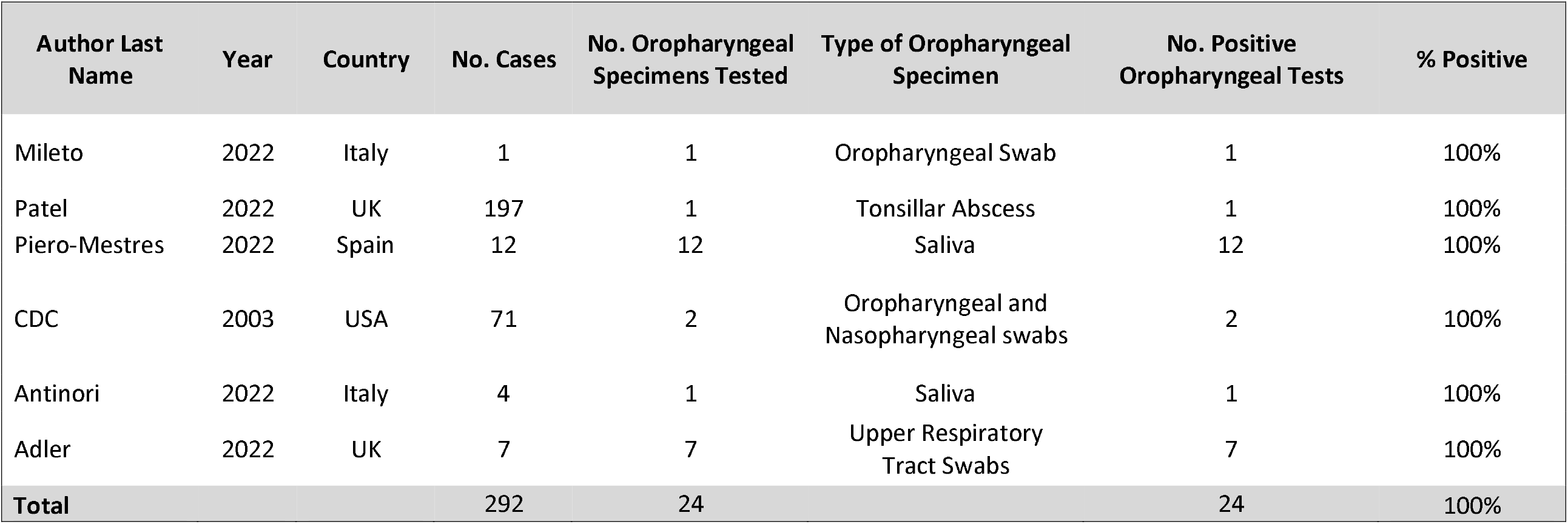
Results of a Systematic Rapid Review Reporting Positivity Among Saliva-Based Tests for Human Monkeypox.

## Discussion

We report the performance and use of a saliva-based PCR test for Human Monkeypox. We supplemented our report of the assay performance with a systematic rapid review of the literature to summarize the performance of saliva-based tests compared to lesion swabs.

Laboratory analysis demonstrated strong agreement between the saliva-based test and known positive and negative specimens. Based on genetic sequence analysis strain from published sequences, all current circulating infections should be detected by the assay. Further genomic analysis from clinical samples will be important to confirm those results *in vivo*. Worth noting, the assay was not specific for Human Monkeypox, but detected several orthopoxvirus subspecies. However, since there is only one clinically relevant outbreak of orthopoxvirus species currently. That lack of specificity is unlikely to be clinically meaningful.

When implemented into clinical practice, 22 cases of Human Monkeypox infection were diagnosed. Importantly, there were no lesion swabs collected simultaneously, thus we cannot comment on the clinical sensitivity and specificity of the assay in those settings. However, among those with positive saliva tests, 16 had clinical disease, one was asymptomatic, and four did not have a rash or lesions at the time of testing. Those findings are of particular importance given the potential utility of saliva-based tests to diagnose Human Monkeypox earlier in the time course of the illness than lesion-based tests.

One report from Belgium identified asymptomatic cases via rectal testing (12). Additionally, a pre-print report from the Democratic Republic of the Congo reported detection of Human Monkeypox DNA from a throat swab of individuals with prodromal symptoms (13). Thus, oral fluid and/or saliva-based tests may have the potential for earlier detection of cases. Earlier case identification would likely result in earlier isolation reducing infectiousness and earlier treatment to reduce the development of infectious lesions. Further work should directly compare the performance of saliva-based tests to lesion swabs in various stages of infection.

One additional consideration beyond earlier detection is the concern that if the virus can be detected prior to lesion development, it may also be transmittable prior to lesion development. Prior work has similarly suggested that viral shedding at various anatomic sites may contribute to transmission. Delayed viral detection may reflect protracted infectiousness (10). In addition, from prior outbreaks of Human Monkeypox human-to-human transmission through respiratory droplets has been documented among a small subset of cases (2).

Two further benefits of saliva-based testing are worth considering. The current outbreak has consistently presented with anogenital lesions (10, 14, 15). Rapid and accessible testing of anogenital lesions may be more challenging than saliva-based tests, given that patients will require privacy to collect anogenital specimens, in contrast to walk-up or drive-through saliva testing centers for SARS-CoV-2. Further, adapting SARS-CoV-2 testing sites for Human Monkeypox saliva testing will rapidly expand testing accessibility and capacity across the country. Beyond convenience, however, the predominance of anogenital lesions in conjunction with data from contact tracing efforts have strongly suggested transmission through sexual contact (4, 5), particularly among men who have sex with men (4). Urgent work is needed to control the outbreak, and saliva-based tests may be a crucial component of those efforts.

One challenge in understanding the performance of saliva-based tests is the heterogeneity with which such tests have been used in published case series (5). Our systematic rapid review, however, demonstrated 100% agreement with lesion PCR testing (5, 10, 16-19). That performance may not reflect the true performance of saliva-based tests, however, given the small overall sample size and that all included studies were case series. From one large study not included in our review as there was no denominator for the number of saliva-based tests from which we could calculate the percent agreement with lesion testing, false negative tests were reported compared to PCR testing of seminal fluid (4). Thus, an accurate and precise estimate of saliva-based test performance in comparison to PCR of lesion swabs is urgently needed.

Our study has several limitations. Regarding the clinical sensitivity of the assay, robust measures could not be assessed given the absence of a comparator gold-standard test. The clinical accuracy of saliva-based tests for Human Monkeypox will be determined by comparisons of the saliva-based test results to other reference standards of infection (i.e., positive viral lesion tests or serological conversion). With regards to the systematic rapid review, the quality of the studies included was poor, and heterogeneous, thus again a definitive determination of the performance of saliva-based tests was not possible. Thus, our findings should be viewed as a call to action in the development and comparison of saliva-based specimen testing.

## Conclusion

We report the analytic performance and use of a saliva-based PCR test for Human Monkeypox. Supplementing that report, we systematically reviewed the literature for all reports of saliva-based tests for Human Monkeypox. Our findings provide evidence that saliva-based tests may be a viable testing method for Human Monkeypox and may identify cases earlier than lesion-based tests. Further evaluation of the performance of saliva-based assays for Human Monkeypox is urgently needed.

## Data Availability

Data are available upon request

## Acknowledgements and Funding

This research was supported in part by a gift to the Keck School of Medicine of the University of Southern California by the W.M. Keck Foundation

