## Supplement 1 for "Laboratory Validation and Clinical Implementation of Human Monkeypox Saliva-Based Tests"

Analysis is based on NCBI complete genomes.

Below is listed the isolates from 2022. All of them are detected by our assay.

**>**ON631963.1 |Monkeypox virus isolate MPxV/VIDRL01/2022, complete genome
**>**ON637938.1 |Monkeypox virus isolate MPXV/Germany/2022/RKI01, complete genome
**>**ON637939.1 |Monkeypox virus isolate MPXV/Germany/2022/RKI02, complete genome
**>**ON631241.1 |Monkeypox virus isolate 2022/2 SLO, complete genome
**>**ON627808.1 |Monkeypox virus isolate MPX/human/USA/UT-UPHL-82200022/2022, complete genome
**>**ON619835.1 |Monkeypox virus isolate MPXV_UK_2022_1, complete genome
**>**ON619836.1 |Monkeypox virus isolate MPXV_UK_2022_2, complete genome
**>**ON619837.1 |Monkeypox virus isolate MPXV_UK_2022_3, complete genome
**>**ON619838.1 |Monkeypox virus isolate MPXV_UK_2022_4, complete genome
**>**ON622712.1 |Monkeypox virus isolate MPX/UZ_REGA_1/Belgium/2022, complete genome
**>**ON622713.1 |Monkeypox virus isolate MPX/UZ_REGA_2/Belgium/2022, complete genome
**>**ON622718.1 |Monkeypox virus isolate MPXV/ES0001/HUGTiP/2022, partial genome
**>**ON622720.1 |Monkeypox virus isolate MPXV-CH-38156923/2022, partial genome
**>**ON622721.1 |Monkeypox virus isolate MPXV_1_IT_Milan_2022, partial genome
**>**ON622722.1 |Monkeypox virus isolate MPXV_FR_HCL0001_2022, complete genome
**>**ON609725.2 |Monkeypox virus isolate SLO, complete genome
**>**ON614676.1 |Monkeypox virus isolate INMI-Pt1, partial genome
**>**ON615424.1 |Monkeypox virus isolate MPXV_2022_NL001, partial genome
**>**ON602722.1 |Monkeypox virus isolate MPXV_FRA_2022_TLS67, complete genome
**>**ON585029.1 |Monkeypox virus isolate Monkeypox/PT0001/2022, partial genome
**>**ON585030.1 |Monkeypox virus isolate Monkeypox/PT0002/2022, partial genome
**>**ON585031.1 |Monkeypox virus isolate Monkeypox/PT0003/2022, complete genome
**>**ON585032.1 |Monkeypox virus isolate Monkeypox/PT0004/2022, complete genome
**>**ON585033.1 |Monkeypox virus isolate Monkeypox/PT0006/2022, complete genome
**>**ON585034.1 |Monkeypox virus isolate Monkeypox/PT0007/2022, complete genome
**>**ON585035.1 |Monkeypox virus isolate Monkeypox/PT0009/2022, complete genome
**>**ON585036.1 |Monkeypox virus isolate Monkeypox/PT0010/2022, partial genome
**>**ON585037.1 |Monkeypox virus isolate Monkeypox/PT0005/2022, complete genome
**>**ON585038.1 |Monkeypox virus isolate Monkeypox/PT0008/2022, complete genome
**>**ON595760.2 |Monkeypox virus isolate MPXV-CH-38134631/2022, partial genome
**>**ON568298.1 |Monkeypox virus isolate MPXV-BY-IMB25241, complete genome
**>**ON563414.3 |Monkeypox virus isolate MPXV_USA_2022_MA001, complete genome

The list of previous strains matching the assay including Congo basin and West Africa

AF380138.1_Monkeypox_virus_strain_Zaire-96-I-16
AY603973.1_Monkeypox_virus_strain_MPXV-WRAIR7-61
AY741551.1_Monkeypox_virus_isolate_Sierra_Leone
AY753185.1_Monkeypox_virus_strain_COP-58
DQ011153.1_Monkeypox_virus_strain_USA_2003_044
DQ011154.1_Monkeypox_virus_strain_Congo_2003_358
DQ011155.1_Monkeypox_virus_strain_Zaire_1979-005
DQ011156.1_Monkeypox_virus_strain_Liberia_1970_184
DQ011157.1_Monkeypox_virus_strain_USA_2003_039
HM172544.1_Monkeypox_virus_strain_Zaire_1979-005
HQ857562.1_Monkeypox_virus_strain_V79-I-005
HQ857563.1_Monkeypox_virus_strain_D14L_knockout
JX878407.1_Monkeypox_virus_isolate_DRC_06-0950
JX878408.1_Monkeypox_virus_isolate_DRC_06-0970
JX878417.1_Monkeypox_virus_isolate_DRC_07-0104
JX878418.1_Monkeypox_virus_isolate_DRC_07-0120
JX878419.1_Monkeypox_virus_isolate_DRC_07-0275
JX878420.1_Monkeypox_virus_isolate_DRC_07-0283
JX878423.1_Monkeypox_virus_isolate_DRC_07-0337
JX878424.1_Monkeypox_virus_isolate_DRC_07-0338
JX878425.1_Monkeypox_virus_isolate_DRC_07-0354
JX878426.1_Monkeypox_virus_isolate_DRC_07-0450
JX878429.1_Monkeypox_virus_isolate_DRC_07-0662
KC257459.1_Monkeypox_virus_strain_Sudan_2005_01
KC257460.1_Monkeypox_virus_strain_DRC_Yandongi_1985
KJ642612.1_Monkeypox_virus_strain_Ikubi
KJ642613.1_Monkeypox_virus_strain_Congo_8
KJ642614.1_Monkeypox_virus_strain_UTC
KJ642615.1_Monkeypox_virus_strain_W-Nigeria
KJ642616.1_Monkeypox_virus_strain_PCH
KJ642617.1_Monkeypox_virus_strain_Nigeria-SE-1971
KJ642618.1_Monkeypox_virus_strain_Cameroon-1990
KJ642619.1_Monkeypox_virus_strain_Gabon-1988
KP849469.1_Monkeypox_virus_isolate_Boende_DRC_2008
KP849470.1_Monkeypox_virus_isolate_Cote_d'Ivoire_1971
KP849471.1_Monkeypox_virus_isolate_Yambuku_DRC_1985
MK783028.1_UNVERIFIED:_Monkeypox_virus_strain_3019
MK783029.1_UNVERIFIED:_Monkeypox_virus_strain_3029
MK783030.1_UNVERIFIED:_Monkeypox_virus_strain_3025
MK783031.1_UNVERIFIED:_Monkeypox_virus_strain_3020
MK783032.1_UNVERIFIED:_Monkeypox_virus_strain_3030
MN346690.1_UNVERIFIED:_Monkeypox_virus_isolate_MPXV_TNP_2017_North_Bic
MN346692.1_UNVERIFIED:_Monkeypox_virus_isolate_MPXV_TNP_2017_North_Mama
MN346693.1_UNVERIFIED:_Monkeypox_virus_isolate_MPXV_TNP_2017_North_Ponan
MN346694.1_UNVERIFIED:_Monkeypox_virus_isolate_MPXV_TNP_2017_North_Saro
MN346695.1_UNVERIFIED:_Monkeypox_virus_isolate_MPXV_TNP_2017_North_Sidonie
MN346696.1_UNVERIFIED:_Monkeypox_virus_isolate_MPXV_TNP_2017_North_Surprise_1
MN346698.1_UNVERIFIED:_Monkeypox_virus_isolate_MPXV_TNP_2017_South_Pushkin
MN346699.1_UNVERIFIED:_Monkeypox_virus_isolate_MPXV_TNP_2017_South_Ravel_1
MN346700.1_UNVERIFIED:_Monkeypox_virus_isolate_MPXV_TNP_2017_South_Ravel_2
MN346702.1_UNVERIFIED:_Monkeypox_virus_isolate_MPXV_TNP_2018_East_Paddy
MN648051.1_Monkeypox_virus_strain_Israel_2018
MT903337.1_Monkeypox_virus_isolate_MPXV-M2940_FCT
MT903338.1_Monkeypox_virus_isolate_MPXV-M2957_Lagos
MT903339.1_Monkeypox_virus_isolate_MPXV-M3021_Delta
MT903340.1_Monkeypox_virus_isolate_MPXV-M5312_HM12_Rivers
MT903341.1_Monkeypox_virus_isolate_MPXV-M5320_M15_Bayelsa
MT903342.1_Monkeypox_virus_isolate_MPXV-Singapore
MT903343.1_Monkeypox_virus_isolate_MPXV-UK_P1
MT903344.1_Monkeypox_virus_isolate_MPXV-UK_P2
MT903345.1_Monkeypox_virus_isolate_MPXV-UK_P3
MT903346.1_Monkeypox_virus_isolate_MPXV-USA2003_099_Gambian_Rat
MT903347.1_Monkeypox_virus_isolate_MPXV-USA2003_099_Dormouse
MT903348.1_Monkeypox_virus_isolate_MPXV-USA2003_099_Rope_Squirrel
NC_003310.1_Monkeypox_virus_Zaire-96-I-16
