## Supplement 2 for "Laboratory Validation and Clinical Implementation of Human Monkeypox Saliva-Based Tests"

### Spectrum Saliva Sample Collection Guide

*****Patient cannot eat, drink, chew gum, or smoke for at least 30 minutes before spitting into the saliva collection tube.*****

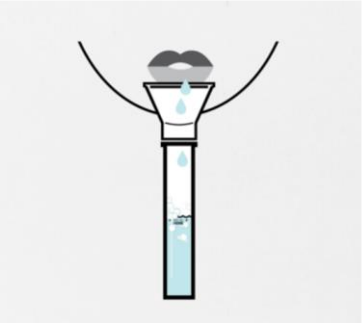

1. Fill tube with saliva (not including bubbles) to the black wavy line. DO NOT OVERFILL.

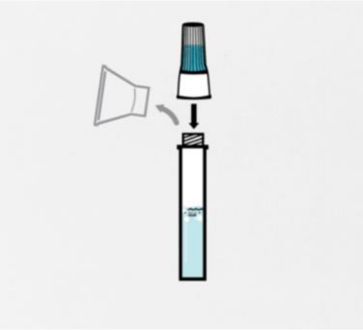

2. Remove the funnel from the tube. Screw on the enclosed cap TIGHTLY to release the solution that will stabilize the DNA in your saliva.

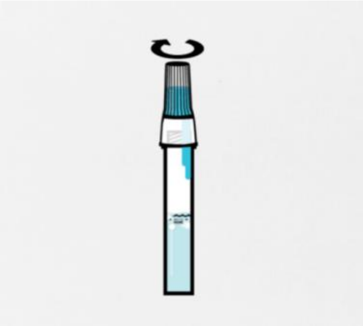

3. Firmly screw cap down to release solution and seal tube.
    a. You will know it works when the blue solution from the cap is released into the tube. Firmly tighten the cap to assure the cap and tube are completely sealed.

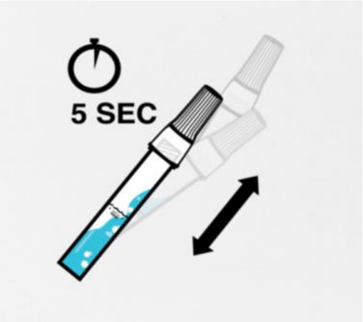


4. Shake the tube for at least five seconds.
    a. This will ensure your sample mixes thoroughly with the stabilizing solution.
