## Supplement 3 for "Laboratory Validation and Clinical Implementation of Human Monkeypox Saliva-Based Tests"

### Analysis of Monkeypox Virus with RT-qPCR Validation Data

### 8. Verification and Validation procedures and results

#### 8.1. Sample preparation for the MonkeyPox verification testing

1. **Prepare Serial Dilutions according to the table Below.**

**Table 10.** Serial dilution of BEI MonkeyPox virus DNA which was used as PC- QPCR for Assay Verification was carried.

| Serial Dilution | Stock/Dilution Volume (uL) | NFW (uL) | Final Volume (uL) | Concentration copies/uL | Notes |
|---|---|---|---|---|---|
| 1 | 9 | 26 | 35 | 1000 | 9uL of Working Stock |
| 2 | 15 | 15 | 30 | 500 | 15uL of Dilution 1 |
| 3 | 15 | 15 | 30 | 250 | 15uL of Dilution 2 |
| 4 | 15 | 15 | 30 | 125 | 15uL of Dilution 3 |
| 5 | 15 | 15 | 30 | 62.5 | 15uL of Dilution 4 |
| 6 | 15 | 15 | 30 | 31.25 | 15uL of Dilution 5 |
| 7 | 15 | 15 | 30 | 15.625 | 15uL of Dilution 6 |

##### Results: LOD for the PC verification using BEI control DNA

| Well Position | Sample | Target | Reporter | Amp Status | Cq | Cq Confidence |
|---|---|---|---|---|---|---|
| A1 | 1000-1 | MPXV | FAM | Amp | 28.45 | 0.89 |
| A1 | 1000-1 | RNaseP | VIC | Amp | 27.21 | 0.99 |
| A2 | 1000-2 | MPXV | FAM | Amp | 28.83 | 0.99 |
| A2 | 1000-2 | RNaseP | VIC | Amp | 27.72 | 0.99 |
| A3 | 1000-3 | MPXV | FAM | Amp | 28.71 | 0.99 |
| A3 | 1000-3 | RNaseP | VIC | Amp | 27.61 | 0.99 |
| B1 | 500-1 | MPXV | FAM | Amp | 29.52 | 0.95 |
| B1 | 500-1 | RNaseP | VIC | Amp | 27.93 | 0.99 |
| B2 | 500-2 | MPXV | FAM | Amp | 29.74 | 0.99 |
| B2 | 500-2 | RNaseP | VIC | Amp | 28.37 | 0.99 |
| B3 | 500-3 | MPXV | FAM | Amp | 29.71 | 0.99 |
| B3 | 500-3 | RNaseP | VIC | Amp | 29.09 | 0.98 |
| C1 | 250-1 | MPXV | FAM | Amp | 27.21 | 0.18 |
| C1 | 250-1 | RNaseP | VIC | Amp | 27.21 | 0.66 |
| C2 | 250-2 | MPXV | FAM | Amp | 29.88 | 0.63 |
| C2 | 250-2 | RNaseP | VIC | Amp | 29.07 | 0.99 |
| C3 | 250-3 | MPXV | FAM | Amp | 30.31 | 0.99 |
| C3 | 250-3 | RNaseP | VIC | Amp | 29.47 | 0.99 |
| D1 | 125-1 | MPXV | FAM | Amp | 31.88 | 0.99 |
| D1 | 125-1 | RNaseP | VIC | Amp | 31.06 | 0.99 |
| D2 | 125-2 | MPXV | FAM | Amp | 30.31 | 0.78 |
| D2 | 125-2 | RNaseP | VIC | Amp | 30.60 | 0.99 |

| | | | | | | |
|---|---|---|---|---|---|---|
| D3 | 125-3 | MPXV | FAM | Amp | 31.68 | 0.99 |
| D3 | 125-3 | RNaseP | VIC | Amp | 31.19 | 0.99 |
| E1 | 62.5-1 | MPXV | FAM | Amp | 31.24 | 0.93 |
| E1 | 62.5-1 | RNaseP | VIC | Amp | 31.66 | 0.99 |
| E2 | 62.5-2 | MPXV | FAM | Amp | 32.77 | 0.98 |
| E2 | 62.5-2 | RNaseP | VIC | Amp | 31.37 | 0.99 |
| E3 | 62.5-3 | MPXV | FAM | No Amp | Undetermined | 0.00 |
| E3 | 62.5-3 | RNaseP | VIC | No Amp | Undetermined | 0.00 |
| F1 | 31.25-1 | MPXV | FAM | Amp | 31.62 | 0.72 |
| F1 | 31.25-1 | RNaseP | VIC | Amp | 32.51 | 0.98 |
| F2 | 31.25-2 | MPXV | FAM | Inconclusive | 34.11 | 0.99 |
| F2 | 31.25-2 | RNaseP | VIC | Amp | 33.82 | 0.99 |
| F3 | 31.25-3 | MPXV | FAM | Amp | 33.86 | 0.98 |
| F3 | 31.25-3 | RNaseP | VIC | Amp | 32.53 | 0.99 |
| G1 | 15.625-1 | MPXV | FAM | No Amp | Undetermined | 0.00 |
| G1 | 15.625-1 | RNaseP | VIC | Amp | 33.88 | 0.99 |
| G2 | 15.625-2 | MPXV | FAM | Inconclusive | 35.14 | 0.97 |
| G2 | 15.625-2 | RNaseP | VIC | Amp | 33.97 | 0.98 |
| G3 | 15.625-3 | MPXV | FAM | Inconclusive | 35.01 | 0.97 |
| G3 | 15.625-3 | RNaseP | VIC | Amp | 33.84 | 0.99 |
| H1 | NEC | MPXV | FAM | No Amp | Undetermined | 0.00 |
| H1 | NEC | RNaseP | VIC | No Amp | Undetermined | 0.00 |
| H2 | NEC2 | MPXV | FAM | No Amp | Undetermined | 0.00 |
| H2 | NEC2 | RNaseP | VIC | No Amp | Undetermined | 0.00 |
| H10 | NTC | MPXV | FAM | Inconclusive | 36.35 | 0.00 |
| H10 | NTC | RNaseP | VIC | Amp | 36.35 | 0.95 |
| H11 | NTC | MPXV | FAM | No Amp | Undetermined | 0.00 |
| H11 | NTC | RNaseP | VIC | Amp | 35.97 | 0.98 |
| H12 | NTC | MPXV | FAM | No Amp | Undetermined | 0.00 |
| H12 | NTC | RNaseP | VIC | Amp | 37.04 | 0.94 |

**Patient 1 and 2 verification using the LA and NY county confirm sample**

**Samples were collected using the saliva spectrum and specimax saliva kit and run.**

| | Well Position | Sample | Target | Reporter | Amp Status | Cq | Cq Confidence |
|---|---|---|---|---|---|---|---|
| Patient 1 (Sal-Spectrum kit, MAX-Specimax kit) | A1 | NEC | MPXV | FAM | No Amp | Undetermined | 0.00 |
| | A1 | NEC | RNaseP | VIC | Inconclusive | 37.70 | 0.95 |
| | D1 | SAL1 | MPXV | FAM | Amp | 31.08 | 0.96 |
| | D1 | SAL1 | RNaseP | VIC | Amp | 12.94 | 0.88 |
| | D2 | SAL2 | MPXV | FAM | Amp | 33.24 | 0.68 |
| | D2 | SAL2 | RNaseP | VIC | Amp | 13.33 | 0.86 |
| | D3 | SAL3 | MPXV | FAM | Amp | 31.61 | 0.97 |
| | D3 | SAL3 | RNaseP | VIC | Amp | 12.76 | 0.87 |
| | D4 | SAL4 | MPXV | FAM | Amp | 29.94 | 0.97 |
| | D4 | SAL4 | RNaseP | VIC | Amp | 11.98 | 0.89 |
| | D5 | SAL5 | MPXV | FAM | Amp | 30.40 | 0.98 |
| | D5 | SAL5 | RNaseP | VIC | Amp | 12.41 | 0.89 |
| | D6 | SAL6 | MPXV | FAM | Amp | 30.94 | 0.98 |
| | D6 | SAL6 | RNaseP | VIC | Amp | 12.37 | 0.86 |
| | D7 | SAL7 | MPXV | FAM | Amp | 32.83 | 0.85 |
| | D7 | SAL7 | RNaseP | VIC | Amp | 12.90 | 0.82 |
| | D8 | SAL8 | MPXV | FAM | Amp | 30.10 | 0.99 |
| | D8 | SAL8 | RNaseP | VIC | Amp | 13.16 | 0.87 |
| | D9 | SAL9 | MPXV | FAM | Amp | 32.76 | 0.78 |
| | D9 | SAL9 | RNaseP | VIC | Amp | 13.36 | 0.57 |

| | | | | | | | |
|---|---|---|---|---|---|---|---|
| | E1 | MAX1 | MPXV | FAM | Amp | 31.87 | 0.99 |
| | E1 | MAX1 | RNaseP | VIC | Amp | 17.96 | 0.87 |
| | E2 | MAX2 | MPXV | FAM | Amp | 29.75 | 0.97 |
| | E2 | MAX2 | RNaseP | VIC | Amp | 18.57 | 0.94 |
| | E3 | MAX3 | MPXV | FAM | Amp | 31.23 | 0.99 |
| | E3 | MAX3 | RNaseP | VIC | Amp | 13.24 | 0.84 |
| | E4 | MAX4 | MPXV | FAM | Amp | 30.32 | 0.99 |
| | E4 | MAX4 | RNaseP | VIC | Amp | 14.88 | 0.84 |
| | E5 | MAX5 | MPXV | FAM | Amp | 31.60 | 0.98 |
| | E5 | MAX5 | RNaseP | VIC | Amp | 15.15 | 0.85 |
| | E6 | MAX6 | MPXV | FAM | Amp | 30.79 | 0.99 |
| | E6 | MAX6 | RNaseP | VIC | Amp | 15.40 | 0.78 |
| | E7 | MAX7 | MPXV | FAM | Amp | 31.81 | 0.97 |
| | E7 | MAX7 | RNaseP | VIC | Amp | 15.94 | 0.83 |
| | E8 | MAX8 | MPXV | FAM | Amp | 30.91 | 0.99 |
| | E8 | MAX8 | RNaseP | VIC | Amp | 16.90 | 0.84 |
| | E9 | MAX9 | MPXV | FAM | Amp | 34.34 | 0.87 |
| | E9 | MAX9 | RNaseP | VIC | Amp | 19.84 | 0.89 |
| | P24 | NTC | MPXV | FAM | Inconclusive | 36.54 | 0.00 |
| | P24 | NTC | RNaseP | VIC | Inconclusive | 36.62 | 0.94 |
| Patient 2 (sal-spectrum saliva kit) | A13 | NEC | MPXV | FAM | No Amp | Undetermined | 0.00 |
| | A13 | NEC | RNaseP | VIC | Inconclusive | 39.23 | 0.00 |
| | A14 | Sal1 | MPXV | FAM | Amp | 23.89 | 0.90 |
| | A14 | Sal1 | RNaseP | VIC | Amp | 16.81 | 0.78 |
| | A15 | Sal2 | MPXV | FAM | Amp | 25.44 | 0.98 |
| | A15 | Sal2 | RNaseP | VIC | Amp | 16.42 | 0.83 |
| | A16 | Sal3 | MPXV | FAM | Amp | 19.65 | 0.17 |
| | A16 | Sal3 | RNaseP | VIC | Amp | 12.95 | 0.77 |
| | A17 | Sal4 | MPXV | FAM | Amp | 23.50 | 0.98 |
| | A17 | Sal4 | RNaseP | VIC | Amp | 17.15 | 0.80 |
| | A18 | Sal5 | MPXV | FAM | Amp | 21.43 | 0.98 |
| | A18 | Sal5 | RNaseP | VIC | Amp | 16.30 | 0.87 |
| | A19 | Sal6 | MPXV | FAM | Amp | 15.11 | 0.04 |
| | A19 | Sal6 | RNaseP | VIC | Amp | 12.99 | 0.63 |
| | A20 | Sal7 | MPXV | FAM | Amp | 21.32 | 0.57 |
| | A20 | Sal7 | RNaseP | VIC | Amp | 16.75 | 0.85 |
| | A21 | Sal8 | MPXV | FAM | Amp | 22.28 | 0.89 |
| | A21 | Sal8 | RNaseP | VIC | Amp | 17.17 | 0.89 |
| | E12 | NTC-1 | MPXV | FAM | No Amp | Undetermined | 0.00 |
| | E12 | NTC-1 | RNaseP | VIC | Amp | 36.04 | 0.97 |
| | F12 | NTC-2 | MPXV | FAM | Inconclusive | 35.72 | 0.02 |
| | F12 | NTC-2 | RNaseP | VIC | Inconclusive | 37.08 | 0.96 |
| | G12 | NTC-3 | MPXV | FAM | No Amp | Undetermined | 0.00 |
| | G12 | NTC-3 | RNaseP | VIC | Amp | 35.00 | 0.97 |
| | H12 | PC | MPXV | FAM | Amp | 26.24 | 0.70 |
| | H12 | PC | RNaseP | VIC | Amp | 25.35 | 0.92 |

BEI control DNA will be used as PC for the studies.

#### 8.2. Limit of Detection – Analytical Sensitivity

Limit of detection (LoD) studies was established by a 2-fold dilution series with pooled Negative saliva specimens' replicates with Thermo MonkeyPox virus DNA Control. To confirm the LoD, 20 replicates for the virus from the LoD determined in the preliminary range finding study were tested.

Acceptance criteria for the LoD study:

1. Preliminary LoD: The lowest concentration that gives MPXV positive results 100% of the time within the dilution series.

2. Final LoD: The lowest concentration at which ≥95% of the replicates (i.e. 19/20 for virus) is positive.

| Viral Single-use Stock | | Working Stock Concentration (copies/uL) | Volume of Dilution Stock (uL) | Volume of NFW |
|---|---|---|---|---|
| Thermo Monkeypox virus DNA Control | | 1000 | 5 | 495 |
| **Serial Dilution** | **Stock/Dilution Volume (ul)** | **Negative saliva matrix(ul)** | **Final Volume (uL)** | **Concentration copies/ml** |
| 1 | 6 | 2996 | 1500 | 2000 |
| 2 | 1500 | 1500 | 1500 | 1000 |
| 3 | 1500 | 1500 | 1500 | 500 |
| 4 | 1500 | 1500 | 1500 | 250 |
| 5 | 1500 | 1500 | 1500 | 125 |
| 6 | 1500 | 1500 | 1500 | 62.5 |
| 7 | 1500 | 1500 | 1500 | 31 |

**Results:**

**Table 12: Preliminary LoD**

| Preliminary LoD MonkeyPox(MPXV) | | | | | | | |
|---|---|---|---|---|---|---|---|
| **Copies/mL** | **Replicates** | **MPXV Gene Ct** | **AMP Status** | **RNaseP ct** | **Amp Status** | **Interpretation** | **% Viral Positivity** |
| MPXV-0 | 1 | Undetermined | No Amp | 16.17 | Amp | Negative MPXV | 0% |
| | 2 | Undetermined | No Amp | 15.96 | Amp | Negative MPXV | |
| | 3 | Undetermined | No Amp | 15.97 | Amp | Negative MPXV | |
| MPXV-31.25 | 1 | Undetermined | No Amp | 15.87 | Amp | Negative MPXV | 33% |
| | 2 | 35.68 | Amp | 15.70 | Amp | Positive MPXV | |
| | 3 | Undetermined | No Amp | 15.84 | Amp | Negative MPXV | |
| MPXV-62.5 | 1 | 34.07 | Amp | 15.46 | Amp | Positive MPXV | 66% |
| | 2 | 34.11 | Amp | 15.62 | Amp | Positive MPXV | |
| | 3 | Undetermined | No Amp | 15.52 | Amp | Negative MPXV | |
| MPXV-125 | 1 | 36.34 | Amp | 15.85 | Amp | Positive MPXV | 66% |
| | 2 | Undetermined | No Amp | 15.76 | Amp | Negative MPXV | |
| | 3 | 34.78 | Amp | 15.78 | Amp | Positive MPXV | |
| MPXV-250 | 1 | 34.58 | Amp | 15.76 | Amp | Positive MPXV | 100% |
| | 2 | 34.60 | Amp | 15.80 | Amp | Positive MPXV | |
| | 3 | 35.71 | Amp | 15.62 | Amp | Positive MPXV | |
| MPXV-500 | 1 | 34.72 | Amp | 15.84 | Amp | Positive MPXV | 100% |
| | 2 | 32.64 | Amp | 15.89 | Amp | Positive MPXV | |
| | 3 | 34.13 | Amp | 15.82 | Amp | Positive MPXV | |
| | 1 | 33.00 | Amp | 15.65 | Amp | Positive MPXV | 100% |

| | | | | | | | |
|---|---|---|---|---|---|---|---|
| MPXV-1000 | 2 | 32.57 | Amp | 15.79 | Amp | Positive MPXV | |
| | 3 | 33.35 | Amp | 15.81 | Amp | Positive MPXV | |
| MPXV-2000 | 1 | 31.91 | Amp | 15.89 | Amp | Positive MPXV | 100% |
| | 2 | 31.92 | Amp | 15.73 | Amp | Positive MPXV | |
| | 3 | 31.54 | Amp | 15.59 | Amp | Positive MPXV | |
| PC-1 | 1 | 17.75 | Amp | 17.48 | Amp | Positive MPXV | 100% |
| PC-2 | 2 | 28.69 | Amp | 26.54 | Amp | Positive MPXV | 100% |
| PC-3 | 3 | 17.47 | Amp | 17.32 | Amp | Positive MPXV | 100% |
| NTC-1 | 1 | Undetermined | No Amp | | No Amp | Negative MPXV | 0% |
| NTC-2 | 2 | Undetermined | No Amp | | No Amp | Negative MPXV | 0% |
| NTC-3 | 3 | Undetermined | No Amp | Undetermined | No Amp | Negative MPXV | 0% |
| NEC-1 | 1 | Undetermined | No Amp | Undetermined | No Amp | Negative MPXV | 0% |
| NEC-2 | 2 | 39.44 | No Amp | Undetermined | No Amp | Negative MPXV | 0% |
| NEC-3 | 3 | Undetermined | No Amp | Undetermined | No Amp | Negative MPXV | 0% |

PC-Positive control, NTC-Non template control , NEC - Negative extraction control

**Confirmation was run at 500 and 250 copies/ml to confirm the LOD of the assay.**

**Table 13. Confirmatory LoD**

| Serial Dilution | Stock/Dilution Volume(uL) | Saliva Negative matrix(uL) | Final Volume (uL) | Concentration copies/mL | Notes |
|---|---|---|---|---|---|
| 1 | 9 | 17991 | 9000 | 500 | 90uL of working stock |
| 2 | 9000 | 9000 | 18000 | 250 | 9000uL of Dilution 1 |

**Table 14: Confirmatory LoD**

| MPXV Confirmation LoD | | | | | | | |
|---|---|---|---|---|---|---|---|
| **Copies/mL** | **Replicates** | **MPXV Gene Ct** | **AMP Status** | **RNaseP ct** | **Amp Status** | **Interpretation** | **% Viral Positivity** |
| 250 cp/mL | 1 | 32.10 | Amp | 16.98 | Amp | MPXV Positive | 100% |
| | 2 | 31.61 | Amp | 16.90 | Amp | MPXV Positive | |
| | 3 | 33.69 | Amp | 16.75 | Amp | MPXV Positive | |
| | 4 | 33.05 | Amp | 17.05 | Amp | MPXV Positive | |
| | 5 | 31.33 | Amp | 16.86 | Amp | MPXV Positive | |
| | 6 | 31.94 | Amp | 16.87 | Amp | MPXV Positive | |
| | 7 | 31.65 | Amp | 16.79 | Amp | MPXV Positive | |
| | 8 | 31.34 | Amp | 16.96 | Amp | MPXV Positive | |
| | 9 | 32.29 | Amp | 16.96 | Amp | MPXV Positive | |
| | 10 | 32.27 | Amp | 16.98 | Amp | MPXV Positive | |
| | 11 | 32.18 | Amp | 17.10 | Amp | MPXV Positive | |
| | 12 | 32.39 | Amp | 16.91 | Amp | MPXV Positive | |
| | 13 | 30.91 | Amp | 16.90 | Amp | MPXV Positive | |
| | 14 | 31.56 | Amp | 16.92 | Amp | MPXV Positive | |
| | 15 | 32.47 | Amp | 16.87 | Amp | MPXV Positive | |

| | | | | | | | |
|---|---|---|---|---|---|---|---|
| | 16 | 32.29 | Amp | 16.91 | Amp | MPXV Positive | |
| | 17 | 32.50 | Amp | 16.96 | Amp | MPXV Positive | |
| | 18 | 31.52 | Amp | 16.98 | Amp | MPXV Positive | |
| | 19 | 33.51 | Amp | 17.02 | Amp | MPXV Positive | |
| | 20 | 32.83 | Amp | 17.09 | Amp | MPXV Positive | |
| 500 cp/mL | 1 | 30.30 | Amp | 16.98 | Amp | MPXV Positive | 100% |
| | 2 | 31.65 | Amp | 17.17 | Amp | MPXV Positive | |
| | 3 | 30.91 | Amp | 16.99 | Amp | MPXV Positive | |
| | 4 | 30.96 | Amp | 16.99 | Amp | MPXV Positive | |
| | 5 | 32.35 | Amp | 17.34 | Amp | MPXV Positive | |
| | 6 | 32.09 | Amp | 17.08 | Amp | MPXV Positive | |
| | 7 | 31.25 | Amp | 17.19 | Amp | MPXV Positive | |
| | 8 | 30.57 | Amp | 16.97 | Amp | MPXV Positive | |
| | 9 | 31.14 | Amp | 16.91 | Amp | MPXV Positive | |
| | 10 | 31.42 | Amp | 16.86 | Amp | MPXV Positive | |
| | 11 | 31.38 | Amp | 16.95 | Amp | MPXV Positive | |
| | 12 | 31.17 | Amp | 16.91 | Amp | MPXV Positive | |
| | 13 | 30.83 | Amp | 16.94 | Amp | MPXV Positive | |
| | 14 | 30.84 | Amp | 16.91 | Amp | MPXV Positive | |
| | 15 | 31.73 | Amp | 16.90 | Amp | MPXV Positive | |
| | 16 | 31.19 | Amp | 17.04 | Amp | MPXV Positive | |
| | 17 | 31.37 | Amp | 17.19 | Amp | MPXV Positive | |
| | 18 | 31.10 | Amp | 16.89 | Amp | MPXV Positive | |
| | 19 | 31.86 | Amp | 17.33 | Amp | MPXV Positive | |
| | 20 | 31.49 | Amp | 16.98 | Amp | MPXV Positive | |

| **Controls** | **MPXV Gene Ct** | **AMP Status** | **RNaseP ct** | **Amp Status** | **Interpretation** |
|---|---|---|---|---|---|
| Positive Control | 28.45 | Amp | 27.48 | Amp | Pass |
| NEC | Undetermined | Undetermined | 37.19 | Amp | Pass |
| NTC | Undetermined | Undetermined | 36.55 | Amp | Pass |

* Undetermined=UND=Not detected

| **MPXVGene Ct average** | **MPXV Ct STDEV** | **RNaseP Gene ct Average** | **RNaseP Ct STDEV** |
|---|---|---|---|
| 32.17 | 0.73 | 16.94 | 0.09 |

**Conclusion:** A detection rate of 100% was achieved (n=20) at a concentration of 250 copies/ml.

#### 8.3 Precision

Precision is defined as closeness of agreement between independent test results obtained under stipulated conditions. Precision is commonly determined by assessing repeatability (i.e., closeness of agreement between independent test results for the same measurement under the same conditions) and reproducibility (i.e., closeness of agreement between independent test results for the same measurement under changed conditions). Precision can be verified or established by assessing day-to-day, run-to-run, and within-run variation (as well as lot variance) by repeat testing of samples, quality control materials, or calibration materials over time.

Samples consisted of a pool of clinical saliva specimens in collection device which were negative for Monkeypox, with or without a purified viral spike-in to create defined viral concentrations.

At least two different operators and instruments were used on 3 separate days at 3 different concentrations (0.1x LoD, 2x LoD, and 5x LoD) of contrived specimens and one negative specimen in triplicate.

Acceptance criteria for precision:

1. 100% agreement for 2x LoD and 5x LoD, for virus.
2. Results at 0.1x LoD were reported (While all are likely to be negative, some may be positive due to stochastic factors in early PCR cycles or sample heterogeneity).

**Table 15: Precision**

| | | | Precision Day 1 | | | | | Precision Day 2 | | | | | Precision Day 3 | | | | |
|---|---|---|---|---|---|---|---|---|---|---|---|---|---|---|---|---|---|
| Sample | LoD | Replicates | MPXV Gene Ct | AMP Status | RNaseP ct | Amp Status | Interpretation | MPXV Gene Ct | AMP Status | RNaseP ct | Amp Status | Interpretation | MPXV Gene Ct | AMP Status | RNaseP ct | Amp Status | Interpretation |
| MPXV | 5x | 1 | 30.69 | Amp | 16.02 | Amp | MPXV Positive | 33.13 | Amp | 16.33 | Amp | MPXV Positive | 31.25 | Amp | 15.79 | Amp | MPXV Positive |
| MPXV | 5x | 2 | 30.87 | Amp | 16.01 | Amp | MPXV Positive | 32.52 | Amp | 16.21 | Amp | MPXV Positive | 32.21 | Amp | 16.00 | Amp | MPXV Positive |
| MPXV | 5x | 3 | 27.65 | Amp | 15.16 | Amp | MPXV Positive | 32.82 | Amp | 16.07 | Amp | MPXV Positive | 33.40 | Amp | 15.72 | Amp | MPXV Positive |
| MPXV | 2x | 1 | 33.15 | Amp | 15.63 | Amp | MPXV Positive | 34.78 | Amp | 16.54 | Amp | MPXV Positive | 34.53 | Amp | 15.95 | Amp | MPXV Positive |
| MPXV | 2x | 2 | 31.41 | Amp | 15.51 | Amp | MPXV Positive | 35.93 | Amp | 16.20 | Amp | MPXV Positive | 34.43 | Amp | 15.99 | Amp | MPXV Positive |
| MPXV | 2x | 3 | 33.25 | Amp | 15.54 | Amp | MPXV Positive | 34.19 | Amp | 16.19 | Amp | MPXV Positive | 32.82 | Amp | 15.75 | Amp | MPXV Positive |
| MPXV | 0.1x | 1 | UND | No Amp | 15.67 | Amp | MPXV Negative | 34.35 | Amp | 15.97 | Amp | MPXV Positive | UND | No Amp | 15.84 | Amp | MPXV Negative |
| MPXV | 0.1x | 2 | UND | No Amp | 15.52 | Amp | MPXV Negative | UND | No Amp | 16.20 | Amp | MPXV Negative | 36.33 | Amp | 15.67 | Amp | MPXV Positive |
| MPXV | 0.1x | 3 | UND | No Amp | 15.62 | Amp | MPXV Negative | 36.00 | Amp | 15.93 | Amp | MPXV Positive | 36.32 | Amp | 15.80 | Amp | MPXV Positive |
| Neg-0-1 | N/A | 1 | 38.15 | Inconclusive | 15.42 | Amp | MPXV Negative | UND | No Amp | 16.03 | Amp | MPXV Negative | UND | No Amp | 15.88 | Amp | MPXV Negative |
| Neg-0-2 | N/A | 2 | UND | No Amp | 15.65 | Amp | MPXV Negative | UND | No Amp | 16.22 | Amp | MPXV Negative | UND | No Amp | 15.81 | Amp | MPXV Negative |
| Neg-0-3 | N/A | 3 | UND | No Amp | 15.83 | Amp | MPXV Negative | UND | No Amp | 16.00 | Amp | MPXV Negative | UND | No Amp | 15.72 | Amp | MPXV Negative |
| PC | N/A | 1 | 28.27 | Amp | 26.49 | Amp | MPXV Positive | 28.84 | Amp | 26.78 | Amp | MPXV Positive | 27.84 | Amp | 26.30 | Amp | MPXV Positive |
| NEC | N/A | 1 | UND | No Amp | 35.35 | Amp | MPXV Negative | UND | No Amp | 37.30 | Inconclusive | MPXV Negative | UND | No Amp | 34.56 | Amp | MPXV Negative |
| NEC-2 | N/A | 1 | UND | No Amp | 34.89 | Amp | MPXV Negative | UND | No Amp | UND | No Amp | MPXV Negative | UND | No Amp | 35.64 | Amp | MPXV Negative |
| NTC | N/A | 1 | UND | No Amp | 36.58 | Amp | MPXV Negative | UND | No Amp | 37.06 | Inconclusive | MPXV Negative | UND | No Amp | 35.84 | Amp | MPXV Negative |

* Undetermined=UND=Not detected

| | Average CT | | SD | | %CV | |
|---|---|---|---|---|---|---|
| | MPXV Gene Ct | RNaseP ct | MPXV Gene SD | RNaseP SD | MPXV Gene | RNaseP |
| 5X | 31.7 | 16.0 | 1.3 | 0.3 | 4.0 | 1.7 |
| | 31.9 | 16.1 | 0.9 | 0.1 | 2.7 | 0.7 |
| | 31.3 | 15.6 | 3.2 | 0.5 | 10.1 | 2.9 |
| 2X | 34.2 | 16.0 | 0.9 | 0.5 | 2.6 | 2.9 |
| | 33.9 | 15.9 | 2.3 | 0.4 | 6.8 | 2.2 |
| | 33.4 | 15.8 | 0.7 | 0.3 | 2.1 | 2.1 |

**Conclusion**

Overall precision of the Monkeypox virus detection showed an acceptable Ct for all controls over a period of 3 days. Comparing across separate PCR plates on 3 days (inter-run) and within single PCR plates (intra-run), we showed 100% agreement between samples at 5X and 2x LoD and %CV of <20%.

The test performance agreement between the operators and instruments were > 95%. All negative controls were negative.

#### 8.4 Accuracy/Clinical Evaluation

Accuracy is defined as the closeness of agreement between a test result and an accepted reference value. Analytical accuracy was determined by concordance between test results from LA and NY county. Two samples that were confirmed as positive for Monkeypox by the LA and NY county Health Departments were used for this study, in addition to 20 negative retrospectives saliva specimens (n=20). The positive samples were diluted into various concentrations in negative matrix.

This study will be a continued evaluation as we collect more specimens and confirm with other validated laboratories in the future.

The results from these studies are summarized in Table.

**Table 16. Accuracy of assay**

| | | Sample | Target | Reporter | Amp Status | Cq |
|---|---|---|---|---|---|---|
| Patient 1- Positive - Confirmed by Los Angeles county -Serial dilution in Negative saliva matrix- Making Contrived Positive samples. | B1 | P1_Stock-1 | MPXV | FAM | Amp | 28.92 |
| | B1 | P1_Stock-1 | RNaseP | VIC | Amp | 13.25 |
| | B2 | P1_Stock-2 | MPXV | FAM | Amp | 27.52 |
| | B2 | P1_Stock-2 | RNaseP | VIC | Amp | 13.08 |
| | B3 | P1_Stock-3 | MPXV | FAM | Amp | 24.44 |
| | B3 | P1_Stock-3 | RNaseP | VIC | Amp | 13.11 |
| | C1 | P1-D1-1 | MPXV | FAM | Amp | 28.95 |
| | C1 | P1-D1-1 | RNaseP | VIC | Amp | 12.96 |
| | C2 | P1-D1-2 | MPXV | FAM | Amp | 30.30 |
| | C2 | P1-D1-2 | RNaseP | VIC | Amp | 13.00 |
| | C3 | P1-D1-3 | MPXV | FAM | Amp | 28.83 |
| | C3 | P1-D1-3 | RNaseP | VIC | Amp | 12.95 |
| | D1 | P1-D2-1 | MPXV | FAM | Amp | 30.81 |
| | D1 | P1-D2-1 | RNaseP | VIC | Amp | 13.70 |
| | D2 | P1-D2-2 | MPXV | FAM | Amp | 29.76 |
| | D2 | P1-D2-2 | RNaseP | VIC | Amp | 13.49 |
| | D3 | P1-D2-3 | MPXV | FAM | Amp | 31.49 |
| | D3 | P1-D2-3 | RNaseP | VIC | Amp | 13.69 |

| | | | | | | |
|---|---|---|---|---|---|---|
| | E1 | P1-D3-1 | MPXV | FAM | Amp | 31.07 |
| | E1 | P1-D3-1 | RNaseP | VIC | Amp | 14.19 |
| | E2 | P1-D3-2 | MPXV | FAM | Amp | 31.55 |
| | E2 | P1-D3-2 | RNaseP | VIC | Amp | 14.21 |
| | E3 | P1-D3-3 | MPXV | FAM | Amp | 31.74 |
| | E3 | P1-D3-3 | RNaseP | VIC | Amp | 14.17 |
| | F1 | P1-D4-1 | MPXV | FAM | Amp | 29.57 |
| | F1 | P1-D4-1 | RNaseP | VIC | Amp | 14.21 |
| | F2 | P1-D4-2 | MPXV | FAM | Amp | 32.55 |
| | F2 | P1-D4-2 | RNaseP | VIC | Amp | 14.42 |
| | F3 | P1-D4-3 | MPXV | FAM | Amp | 32.61 |
| | F3 | P1-D4-3 | RNaseP | VIC | Amp | 14.43 |
| | G1 | P1-D5-1 | MPXV | FAM | Amp | 34.55 |
| | G1 | P1-D5-1 | RNaseP | VIC | Amp | 14.58 |
| | G2 | P1-D5-2 | MPXV | FAM | No Amp | UND |
| | G2 | P1-D5-2 | RNaseP | VIC | Amp | 14.70 |
| | G3 | P1-D5-3 | MPXV | FAM | Amp | 34.07 |
| | G3 | P1-D5-3 | RNaseP | VIC | Amp | 14.44 |
| | H1 | P1-D6-1 | MPXV | FAM | No Amp | UND |
| | H1 | P1-D6-1 | RNaseP | VIC | Amp | 14.73 |
| | H2 | P1-D6-2 | MPXV | FAM | No Amp | UND |
| | H2 | P1-D6-2 | RNaseP | VIC | Amp | 14.72 |
| | H3 | P1-D6-3 | MPXV | FAM | No Amp | UND |
| | H3 | P1-D6-3 | RNaseP | VIC | Amp | 14.65 |
| | A4 | NEG1 | MPXV | FAM | No Amp | UND |
| | A4 | NEG1 | RNaseP | VIC | Amp | 15.17 |
| | A5 | NEG2 | MPXV | FAM | No Amp | UND |
| | A5 | NEG2 | RNaseP | VIC | Amp | 14.79 |
| | A6 | NEG3 | MPXV | FAM | No Amp | UND |
| | A6 | NEG3 | RNaseP | VIC | Amp | 14.67 |
| | H12 | PC | MPXV | FAM | Amp | 26.52 |
| | H12 | PC | RNaseP | VIC | Amp | 25.03 |
| | A1 | NEC1 | MPXV | FAM | No Amp | UND |
| | A1 | NEC1 | RNaseP | VIC | Amp | 34.97 |
| | A2 | NEC2 | MPXV | FAM | No Amp | UND |
| | A2 | NEC2 | RNaseP | VIC | No Amp | UND |
| | A3 | NEC3 | MPXV | FAM | No Amp | UND |
| | A3 | NEC3 | RNaseP | VIC | Amp | 35.22 |
| | P22 | NTC1 | MPXV | FAM | No Amp | UND |
| | P22 | NTC1 | RNaseP | VIC | No Amp | UND |
| | P23 | NTC2 | MPXV | FAM | Inconclusive | 39.46 |
| | P23 | NTC2 | RNaseP | VIC | No Amp | UND |
| | P24 | NTC3 | MPXV | FAM | Inconclusive | 35.47 |
| | P24 | NTC3 | RNaseP | VIC | No Amp | UND |
| Patient2- Positive - Confirmed by New York county -Serial dilution in Negative saliva matrix- Making Contrived | A13 | P2-Stock-1 | MPXV | FAM | Amp | 19.90 |
| | A13 | P2-Stock-1 | RNaseP | VIC | Amp | 15.54 |
| | A14 | P2-Stock-2 | MPXV | FAM | Amp | 24.43 |
| | A14 | P2-Stock-2 | RNaseP | VIC | Amp | 15.51 |
| | A15 | P2-Stock-3 | MPXV | FAM | Amp | 24.77 |
| | A15 | P2-Stock-3 | RNaseP | VIC | Amp | 15.74 |
| | B13 | P2-D1-1 | MPXV | FAM | Amp | 24.56 |
| | B13 | P2-D1-1 | RNaseP | VIC | Amp | 14.81 |
| | B14 | P2-D1-2 | MPXV | FAM | Amp | 24.85 |
| | B14 | P2-D1-2 | RNaseP | VIC | Amp | 15.05 |

| | | | | | | |
|---|---|---|---|---|---|---|
| Positive samples. | B15 | P2-D1-3 | MPXV | FAM | Amp | 24.84 |
| | B15 | P2-D1-3 | RNaseP | VIC | Amp | 14.84 |
| | C13 | P2-D2-1 | MPXV | FAM | Amp | 25.50 |
| | C13 | P2-D2-1 | RNaseP | VIC | Amp | 14.86 |
| | C14 | P2-D2-2 | MPXV | FAM | Amp | 26.06 |
| | C14 | P2-D2-2 | RNaseP | VIC | Amp | 14.87 |
| | C15 | P2-D2-3 | MPXV | FAM | Amp | 25.79 |
| | C15 | P2-D2-3 | RNaseP | VIC | Amp | 14.79 |
| | D13 | P2-D3-1 | MPXV | FAM | Amp | 26.53 |
| | D13 | P2-D3-1 | RNaseP | VIC | Amp | 14.75 |
| | D14 | P2-D3-2 | MPXV | FAM | Amp | 26.35 |
| | D14 | P2-D3-2 | RNaseP | VIC | Amp | 14.70 |
| | D15 | P2-D3-3 | MPXV | FAM | Amp | 26.76 |
| | D15 | P2-D3-3 | RNaseP | VIC | Amp | 14.81 |
| | E13 | P2-D4-1 | MPXV | FAM | Amp | 27.55 |
| | E13 | P2-D4-1 | RNaseP | VIC | Amp | 14.96 |
| | E14 | P2-D4-2 | MPXV | FAM | Amp | 27.69 |
| | E14 | P2-D4-2 | RNaseP | VIC | Amp | 14.54 |
| | E15 | P2-D4-3 | MPXV | FAM | Amp | 27.81 |
| | E15 | P2-D4-3 | RNaseP | VIC | Amp | 14.70 |
| | F13 | P2-D5-1 | MPXV | FAM | Amp | 28.98 |
| | F13 | P2-D5-1 | RNaseP | VIC | Amp | 14.97 |
| | F14 | P2-D5-2 | MPXV | FAM | Amp | 28.90 |
| | F14 | P2-D5-2 | RNaseP | VIC | Amp | 14.92 |
| | F15 | P2-D5-3 | MPXV | FAM | Amp | 28.66 |
| | F15 | P2-D5-3 | RNaseP | VIC | Amp | 14.89 |
| | A16 | P2-D6-1 | MPXV | FAM | Amp | 29.59 |
| | A16 | P2-D6-1 | RNaseP | VIC | Amp | 14.85 |
| | A17 | P2-D6-2 | MPXV | FAM | Amp | 29.30 |
| | A17 | P2-D6-2 | RNaseP | VIC | Amp | 14.85 |
| | A18 | P2-D6-3 | MPXV | FAM | Amp | 29.07 |
| | A18 | P2-D6-3 | RNaseP | VIC | Amp | 14.98 |
| | B16 | P2-D7-1 | MPXV | FAM | Amp | 30.06 |
| | B16 | P2-D7-1 | RNaseP | VIC | Amp | 14.74 |
| | B17 | P2-D7-2 | MPXV | FAM | Amp | 29.47 |
| | B17 | P2-D7-2 | RNaseP | VIC | Amp | 14.64 |
| | B18 | P2-D7-3 | MPXV | FAM | Amp | 29.97 |
| | B18 | P2-D7-3 | RNaseP | VIC | Amp | 14.58 |
| | C16 | P2-D8-1 | MPXV | FAM | Amp | 20.93 |
| | C16 | P2-D8-1 | RNaseP | VIC | Amp | 13.69 |
| | C17 | P2-D8-2 | MPXV | FAM | Amp | 19.18 |
| | C17 | P2-D8-2 | RNaseP | VIC | Amp | 14.35 |
| | C18 | P2-D8-3 | MPXV | FAM | Amp | 20.50 |
| | C18 | P2-D8-3 | RNaseP | VIC | Amp | 13.81 |
| | D16 | P2-D9-1 | MPXV | FAM | Amp | 24.60 |
| | D16 | P2-D9-1 | RNaseP | VIC | Amp | 13.37 |
| | D17 | P2-D9-2 | MPXV | FAM | Amp | 18.97 |
| | D17 | P2-D9-2 | RNaseP | VIC | Amp | 13.36 |
| | D18 | P2-D9-3 | MPXV | FAM | Amp | 18.35 |
| | D18 | P2-D9-3 | RNaseP | VIC | Amp | 13.57 |
| | E16 | P2-D10-1 | MPXV | FAM | Amp | 21.59 |
| | E16 | P2-D10-1 | RNaseP | VIC | Amp | 13.26 |
| | E17 | P2-D10-2 | MPXV | FAM | Amp | 19.90 |
| | E17 | P2-D10-2 | RNaseP | VIC | Amp | 13.38 |

| | | | | | | |
|---|---|---|---|---|---|---|
| | E18 | P2-D10-3 | MPXV | FAM | Inconclusive | 18.98 |
| | E18 | P2-D10-3 | RNaseP | VIC | Amp | 13.49 |
| | F16 | Neg-2-1 | MPXV | FAM | Inconclusive | 19.38 |
| | F16 | Neg-2-1 | RNaseP | VIC | Amp | 13.43 |
| | F17 | Neg-2-2 | MPXV | FAM | Inconclusive | 21.34 |
| | F17 | Neg-2-2 | RNaseP | VIC | Amp | 13.49 |
| | F18 | Neg-2-3 | MPXV | FAM | No Amp | UND |
| | F18 | Neg-2-3 | RNaseP | VIC | Amp | 13.97 |
| | H12 | PC | MPXV | FAM | Amp | 27.46 |
| | H12 | PC | RNaseP | VIC | Amp | 25.84 |
| | A22 | NEC-1 | MPXV | FAM | No Amp | UND |
| | A22 | NEC-1 | RNaseP | VIC | Inconclusive | 37.69 |
| | A23 | NEC-2 | MPXV | FAM | No Amp | UND |
| | A23 | NEC-2 | RNaseP | VIC | Amp | 35.99 |
| | A24 | NEC-3 | MPXV | FAM | No Amp | UND |
| | A24 | NEC-3 | RNaseP | VIC | Inconclusive | 36.98 |
| | P22 | NTC | MPXV | FAM | No Amp | UND |
| | P22 | NTC | RNaseP | VIC | Amp | 37.44 |
| | P23 | NTC | MPXV | FAM | No Amp | UND |
| | P23 | NTC | RNaseP | VIC | Inconclusive | 38.15 |
| | P24 | NTC | MPXV | FAM | No Amp | UND |
| | P24 | NTC | RNaseP | VIC | No Amp | UND |

* Undetermined=UND=Not detected

**Conclusion**

All positive samples showed 100% concordance at different dilutions (Patient1- 6*3=24 and Patient 2-9*3= 27). Ct values for RNaseP were < 33 for all samples indicating good extraction.

100% of the negative saliva samples showed negative results for Monkeypox virus.

| Sample | Target | Amp Status | Cq |
|---|---|---|---|
| NEC | MPXV | No Amp | Undetermined |
| NEC | RNAse P | Amp | 37.94 |
| POS-1 | MPXV | No Amp | Undetermined |
| POS-1 | RNAse P | Amp | 19.65 |
| POS-2 | MPXV | No Amp | Undetermined |
| POS-2 | RNAse P | Amp | 15.64 |
| POS-3 | MPXV | No Amp | Undetermined |
| POS-3 | RNAse P | Amp | 19.52 |
| POS-4 | MPXV | No Amp | Undetermined |
| POS-4 | RNAse P | Amp | 14.59 |
| POS-5 | MPXV | No Amp | Undetermined |
| POS-5 | RNAse P | Amp | 15.85 |
| POS-6 | MPXV | No Amp | Undetermined |
| POS-6 | RNAse P | Amp | 19.92 |
| POS-7 | MPXV | No Amp | Undetermined |
| POS-7 | RNAse P | Amp | 15.56 |
| POS-8 | MPXV | No Amp | Undetermined |
| POS-8 | RNAse P | Amp | 16.90 |

| | | | |
|---|---|---|---|
| POS-9 | MPXV | No Amp | Undetermined |
| POS-9 | RNAse P | Amp | 15.04 |
| POS-10 | MPXV | No Amp | Undetermined |
| POS-10 | RNAse P | Amp | 17.79 |
| POS-11 | MPXV | No Amp | Undetermined |
| POS-11 | RNAse P | Amp | 17.42 |
| POS-12 | MPXV | No Amp | Undetermined |
| POS-12 | RNAse P | Amp | 17.79 |
| POS-13 | MPXV | No Amp | Undetermined |
| POS-13 | RNAse P | Amp | 23.11 |
| POS-14 | MPXV | No Amp | Undetermined |
| POS-14 | RNAse P | Amp | 15.73 |
| POS-15 | MPXV | No Amp | Undetermined |
| POS-15 | RNAse P | Amp | 19.53 |
| POS-16 | MPXV | No Amp | Undetermined |
| POS-16 | RNAse P | Amp | 22.83 |
| POS-17 | MPXV | No Amp | Undetermined |
| POS-17 | RNAse P | Amp | 16.41 |
| POS-18 | MPXV | No Amp | Undetermined |
| POS-18 | RNAse P | Amp | 20.39 |
| POS-19 | MPXV | No Amp | Undetermined |
| POS-19 | RNAse P | Amp | 15.64 |
| POS-20 | MPXV | No Amp | Undetermined |
| POS-20 | RNAse P | Amp | 18.13 |
| PC | MPXV | Amp | 27.74 |
| PC | RNAse P | Amp | 26.87 |
| NTC | MPXV | No Amp | Undetermined |
| NTC | RNAse P | No Amp | Undetermined |

#### 8.5. Inclusivity (analytical reactivity)

ThermoFisher developed the primers and probe, documented the methodology and presented results of an in silico inclusivity analysis that established the extent to which variation in the Monkeypox genome may impact sensitivity of test performance. ThermoFisher TaqMan assay can detects both the clades of Monkeypox.

Thermo analysis is based on NCBI complete genomes.

Below is listed the isolates from 2022. All of them are detected by the assay.

| | The list of previous strains matching the assay including Congo basin and West Africa |
|---|---|
| >ON631963.1 \|Monkeypox virus isolate MPxV/VIDRL01/2022, complete genome | AF380138.1_Monkeypox_virus_strain_Zaire-96-I-16 |
| >ON637938.1 \|Monkeypox virus isolate MPXV/Germany/2022/RKI01, complete genome | AY603973.1_Monkeypox_virus_strain_MPXV-WRAIR7-61 |
| >ON637939.1 \|Monkeypox virus isolate MPXV/Germany/2022/RKI02, complete genome | AY741551.1_Monkeypox_virus_isolate_Sierra_Leone |
| >ON631241.1 \|Monkeypox virus isolate 2022/2 SLO, complete genome | AY753185.1_Monkeypox_virus_strain_COP-58 |

POS-8
POS-8

| | |
|---|---|
| >ON627808.1 \|Monkeypox virus isolate MPX/human/USA/UT-UPHL-82200022/2022, complete genome | DQ011153.1_Monkeypox_virus_strain_USA_2003_044 |
| >ON619835.1 \|Monkeypox virus isolate MPXV_UK_2022_1, complete genome | DQ011154.1_Monkeypox_virus_strain_Congo_2003_358 |
| >ON619836.1 \|Monkeypox virus isolate MPXV_UK_2022_2, complete genome | DQ011155.1_Monkeypox_virus_strain_Zaire_1979-005 |
| >ON619837.1 \|Monkeypox virus isolate MPXV_UK_2022_3, complete genome | DQ011156.1_Monkeypox_virus_strain_Liberia_1970_184 |
| >ON619838.1 \|Monkeypox virus isolate MPXV_UK_2022_4, complete genome | DQ011157.1_Monkeypox_virus_strain_USA_2003_039 |
| >ON622712.1 \|Monkeypox virus isolate MPX/UZ_REGA_1/Belgium/2022, complete genome | HM172544.1_Monkeypox_virus_strain_Zaire_1979-005 |
| >ON622713.1 \|Monkeypox virus isolate MPX/UZ_REGA_2/Belgium/2022, complete genome | HQ857562.1_Monkeypox_virus_strain_V79-I-005 |
| >ON622718.1 \|Monkeypox virus isolate MPXV/ES0001/HUGTiP/2022, partial genome | HQ857563.1_Monkeypox_virus_strain_D14L_knockout |
| >ON622720.1 \|Monkeypox virus isolate MPXV-CH-38156923/2022, partial genome | JX878407.1_Monkeypox_virus_isolate_DRC_06-0950 |
| >ON622721.1 \|Monkeypox virus isolate MPXV_1_IT_Milan_2022, partial genome | JX878408.1_Monkeypox_virus_isolate_DRC_06-0970 |
| >ON622722.1 \|Monkeypox virus isolate MPXV_FR_HCL0001_2022, complete genome | JX878417.1_Monkeypox_virus_isolate_DRC_07-0104 |
| >ON609725.2 \|Monkeypox virus isolate SLO, complete genome | JX878418.1_Monkeypox_virus_isolate_DRC_07-0120 |
| >ON614676.1 \|Monkeypox virus isolate INMI-Pt1, partial genome | JX878419.1_Monkeypox_virus_isolate_DRC_07-0275 |
| >ON615424.1 \|Monkeypox virus isolate MPXV_2022_NL001, partial genome | JX878420.1_Monkeypox_virus_isolate_DRC_07-0283 |
| >ON602722.1 \|Monkeypox virus isolate MPXV_FRA_2022_TLS67, complete genome | JX878423.1_Monkeypox_virus_isolate_DRC_07-0337 |
| >ON585029.1 \|Monkeypox virus isolate Monkeypox/PT0001/2022, partial genome | JX878424.1_Monkeypox_virus_isolate_DRC_07-0338 |
| >ON585030.1 \|Monkeypox virus isolate Monkeypox/PT0002/2022, partial genome | JX878425.1_Monkeypox_virus_isolate_DRC_07-0354 |
| >ON585031.1 \|Monkeypox virus isolate Monkeypox/PT0003/2022, complete genome | JX878426.1_Monkeypox_virus_isolate_DRC_07-0450 |
| >ON585032.1 \|Monkeypox virus isolate Monkeypox/PT0004/2022, complete genome | JX878429.1_Monkeypox_virus_isolate_DRC_07-0662 |
| >ON585033.1 \|Monkeypox virus isolate Monkeypox/PT0006/2022, complete genome | KC257459.1_Monkeypox_virus_strain_Sudan_2005_01 |
| >ON585034.1 \|Monkeypox virus isolate Monkeypox/PT0007/2022, complete genome | KC257460.1_Monkeypox_virus_strain_DRC_Yandongi_1985 |
| >ON585035.1 \|Monkeypox virus isolate Monkeypox/PT0009/2022, complete genome | KJ642612.1_Monkeypox_virus_strain_Ikubi |
| >ON585036.1 \|Monkeypox virus isolate Monkeypox/PT0010/2022, partial genome | KJ642613.1_Monkeypox_virus_strain_Congo_8 |
| >ON585037.1 \|Monkeypox virus isolate Monkeypox/PT0005/2022, complete genome | KJ642614.1_Monkeypox_virus_strain_UTC |
| >ON585038.1 \|Monkeypox virus isolate Monkeypox/PT0008/2022, complete genome | KJ642615.1_Monkeypox_virus_strain_W-Nigeria |
| >ON595760.2 \|Monkeypox virus isolate MPXV-CH-38134631/2022, partial genome | KJ642616.1_Monkeypox_virus_strain_PCH |
| >ON568298.1 \|Monkeypox virus isolate MPXV-BY-IMB25241, complete genome | KJ642617.1_Monkeypox_virus_strain_Nigeria-SE-1971 |
| >ON563414.3 \|Monkeypox virus isolate MPXV_USA_2022_MA001, complete genome | KJ642618.1_Monkeypox_virus_strain_Cameroon-1990 |

| | |
|---|---|
| MK783028.1_UNVERIFIED:_Monkeypox_virus_strain_3019 | KJ642619.1_Monkeypox_virus_strain_Gabon-1988 |
| MK783029.1_UNVERIFIED:_Monkeypox_virus_strain_3029 | KP849469.1_Monkeypox_virus_isolate_Boende_DRC_2008 |
| MK783030.1_UNVERIFIED:_Monkeypox_virus_strain_3025 | KP849470.1_Monkeypox_virus_isolate_Cote_d'Ivoire_1971 |
| MK783031.1_UNVERIFIED:_Monkeypox_virus_strain_3020 | KP849471.1_Monkeypox_virus_isolate_Yambuku_DRC_1985 |
| MK783032.1_UNVERIFIED:_Monkeypox_virus_strain_3030 | MN648051.1_Monkeypox_virus_strain_Israel_2018 |
| MN346690.1_UNVERIFIED:_Monkeypox_virus_isolate_MPXV_TNP_2017_North_Bic | MT903337.1_Monkeypox_virus_isolate_MPXV-M2940_FCT |
| MN346692.1_UNVERIFIED:_Monkeypox_virus_isolate_MPXV_TNP_2017_North_Mama | MT903338.1_Monkeypox_virus_isolate_MPXV-M2957_Lagos |
| MN346693.1_UNVERIFIED:_Monkeypox_virus_isolate_MPXV_TNP_2017_North_Ponan | MT903339.1_Monkeypox_virus_isolate_MPXV-M3021_Delta |
| MN346694.1_UNVERIFIED:_Monkeypox_virus_isolate_MPXV_TNP_2017_North_Saro | MT903340.1_Monkeypox_virus_isolate_MPXV-M5312_HM12_Rivers |
| MN346695.1_UNVERIFIED:_Monkeypox_virus_isolate_MPXV_TNP_2017_North_Sidonie | MT903341.1_Monkeypox_virus_isolate_MPXV-M5320_M15_Bayelsa |
| MN346696.1_UNVERIFIED:_Monkeypox_virus_isolate_MPXV_TNP_2017_North_Surprise_1 | MT903342.1_Monkeypox_virus_isolate_MPXV-Singapore |
| MN346698.1_UNVERIFIED:_Monkeypox_virus_isolate_MPXV_TNP_2017_South_Pushkin | MT903343.1_Monkeypox_virus_isolate_MPXV-UK_P1 |
| MN346699.1_UNVERIFIED:_Monkeypox_virus_isolate_MPXV_TNP_2017_South_Ravel_1 | MT903344.1_Monkeypox_virus_isolate_MPXV-UK_P2 |
| MN346700.1_UNVERIFIED:_Monkeypox_virus_isolate_MPXV_TNP_2017_South_Ravel_2 | MT903345.1_Monkeypox_virus_isolate_MPXV-UK_P3 |
| MN346702.1_UNVERIFIED:_Monkeypox_virus_isolate_MPXV_TNP_2018_East_Paddy | MT903346.1_Monkeypox_virus_isolate_MPXV-USA2003_099_Gambian_Rat |
| NC_003310.1_Monkeypox_virus_Zaire-96-I-16 | MT903347.1_Monkeypox_virus_isolate_MPXV-USA2003_099_Dormouse |
| | MT903348.1_Monkeypox_virus_isolate_MPXV-USA2003_099_Rope_Squirrel |

**Conclusion:**

Both clades of Monkeypox can be detected using the primer and probe design as shown in the table above.

#### 8.6. Cross contamination

To determine cross-contamination potential on the Kingfisher instrument, a minimum of 20 contrived positive specimens at 5x LoD were run in a checkerboard pattern (Contrived positive specimens containing MPXV plated in every other well)

**Acceptance criteria for cross-contamination:**

1. ≥90% of contrived positive specimens are positive for MPXV and RNaseP as expected.
2. ≥90% of negative samples are negative for MPXV.
    a. Note: If less than 90% of negative specimens yield negative detection, the Kingfisher instrument protocol will be re-evaluated.

**Results**

| Well | Well Position | Sample | Target | Reporter | Amp Status | Cq |
|---|---|---|---|---|---|---|

| | | | | | | |
|---|---|---|---|---|---|---|
| 2 | A2 | NEC-1 | MPXV | FAM | No Amp | Undetermined |
| 2 | A2 | NEC-1 | RNAse P | VIC | Inconclusive | 39.03 |
| 3 | A3 | MPX-5 | MPXV | FAM | Amp | 34.33 |
| 3 | A3 | MPX-5 | RNAse P | VIC | Amp | 16.05 |
| 4 | A4 | NEC-9 | MPXV | FAM | No Amp | Undetermined |
| 4 | A4 | NEC-9 | RNAse P | VIC | Amp | 37.27 |
| 6 | A6 | NEC-13 | MPXV | FAM | No Amp | Undetermined |
| 6 | A6 | NEC-13 | RNAse P | VIC | No Amp | Undetermined |
| 8 | A8 | MPX-17 | MPXV | FAM | Amp | 33.80 |
| 8 | A8 | MPX-17 | RNAse P | VIC | Amp | 15.92 |
| 10 | A10 | NEC-21 | MPXV | FAM | No Amp | Undetermined |
| 10 | A10 | NEC-21 | RNAse P | VIC | No Amp | Undetermined |
| 26 | B2 | MPX-1 | MPXV | FAM | Amp | 33.54 |
| 26 | B2 | MPX-1 | RNAse P | VIC | Amp | 15.91 |
| 27 | B3 | NEC-5 | MPXV | FAM | No Amp | Undetermined |
| 27 | B3 | NEC-5 | RNAse P | VIC | No Amp | Undetermined |
| 28 | B4 | MPX-9 | MPXV | FAM | Amp | 33.00 |
| 28 | B4 | MPX-9 | RNAse P | VIC | Amp | 15.92 |
| 30 | B6 | MPX-13 | MPXV | FAM | Amp | 34.92 |
| 30 | B6 | MPX-13 | RNAse P | VIC | Amp | 15.99 |
| 32 | B8 | NEC-17 | MPXV | FAM | No Amp | Undetermined |
| 32 | B8 | NEC-17 | RNAse P | VIC | Inconclusive | 37.50 |
| 34 | B10 | MPX-21 | MPXV | FAM | Amp | 34.35 |
| 34 | B10 | MPX-21 | RNAse P | VIC | Amp | 15.79 |
| 36 | B12 | NEC1 | MPXV | FAM | Inconclusive | 39.47 |
| 36 | B12 | NEC1 | RNAse P | VIC | No Amp | Undetermined |
| 50 | C2 | NEC-2 | MPXV | FAM | Inconclusive | 38.64 |
| 50 | C2 | NEC-2 | RNAse P | VIC | No Amp | Undetermined |
| 51 | C3 | MPX-6 | MPXV | FAM | Amp | 33.55 |
| 51 | C3 | MPX-6 | RNAse P | VIC | Amp | 15.83 |
| 52 | C4 | NEC-10 | MPXV | FAM | No Amp | Undetermined |
| 52 | C4 | NEC-10 | RNAse P | VIC | No Amp | Undetermined |
| 54 | C6 | NEC-14 | MPXV | FAM | No Amp | Undetermined |
| 54 | C6 | NEC-14 | RNAse P | VIC | No Amp | Undetermined |
| 56 | C8 | MPX-18 | MPXV | FAM | Amp | 33.83 |
| 56 | C8 | MPX-18 | RNAse P | VIC | Amp | 15.85 |
| 58 | C10 | NEC-22 | MPXV | FAM | No Amp | Undetermined |
| 58 | C10 | NEC-22 | RNAse P | VIC | No Amp | Undetermined |
| 60 | C12 | NEC2 | MPXV | FAM | No Amp | Undetermined |
| 60 | C12 | NEC2 | RNAse P | VIC | No Amp | Undetermined |
| 74 | D2 | MPX-2 | MPXV | FAM | Amp | 34.03 |
| 74 | D2 | MPX-2 | RNAse P | VIC | Amp | 16.07 |
| 75 | D3 | NEC-6 | MPXV | FAM | No Amp | Undetermined |
| 75 | D3 | NEC-6 | RNAse P | VIC | Amp | 36.73 |
| 76 | D4 | MPX-10 | MPXV | FAM | Amp | 34.11 |
| 76 | D4 | MPX-10 | RNAse P | VIC | Amp | 15.92 |
| 78 | D6 | MPX-14 | MPXV | FAM | Amp | 34.31 |
| 78 | D6 | MPX-14 | RNAse P | VIC | Amp | 15.87 |
| 80 | D8 | NEC-18 | MPXV | FAM | No Amp | Undetermined |
| 80 | D8 | NEC-18 | RNAse P | VIC | No Amp | Undetermined |
| 82 | D10 | MPX-22 | MPXV | FAM | Amp | 34.22 |
| 82 | D10 | MPX-22 | RNAse P | VIC | Amp | 15.69 |
| 98 | E2 | NEC-3 | MPXV | FAM | No Amp | Undetermined |
| 98 | E2 | NEC-3 | RNAse P | VIC | Amp | 37.07 |

| | | | | | | |
|---|---|---|---|---|---|---|
| 99 | E3 | MPX-7 | MPXV | FAM | Amp | 34.75 |
| 99 | E3 | MPX-7 | RNAse P | VIC | Amp | 15.98 |
| 100 | E4 | NEC-11 | MPXV | FAM | No Amp | Undetermined |
| 100 | E4 | NEC-11 | RNAse P | VIC | Amp | 36.97 |
| 102 | E6 | NEC-15 | MPXV | FAM | No Amp | Undetermined |
| 102 | E6 | NEC-15 | RNAse P | VIC | No Amp | Undetermined |
| 104 | E8 | MPX--19 | MPXV | FAM | Amp | 34.25 |
| 104 | E8 | MPX--19 | RNAse P | VIC | Amp | 15.81 |
| 106 | E10 | NEC-23 | MPXV | FAM | No Amp | Undetermined |
| 106 | E10 | NEC-23 | RNAse P | VIC | Inconclusive | 38.92 |
| 122 | F2 | MPX-3 | MPXV | FAM | Amp | 33.94 |
| 122 | F2 | MPX-3 | RNAse P | VIC | Amp | 15.92 |
| 123 | F3 | NEC-7 | MPXV | FAM | No Amp | Undetermined |
| 123 | F3 | NEC-7 | RNAse P | VIC | No Amp | Undetermined |
| 124 | F4 | MPX-11 | MPXV | FAM | Amp | 33.48 |
| 124 | F4 | MPX-11 | RNAse P | VIC | Amp | 15.83 |
| 126 | F6 | MPX-15 | MPXV | FAM | Amp | 34.86 |
| 126 | F6 | MPX-15 | RNAse P | VIC | Amp | 15.78 |
| 128 | F8 | NEC-19 | MPXV | FAM | No Amp | Undetermined |
| 128 | F8 | NEC-19 | RNAse P | VIC | Inconclusive | 37.83 |
| 130 | F10 | MPX-23 | MPXV | FAM | Amp | 34.10 |
| 130 | F10 | MPX-23 | RNAse P | VIC | Amp | 15.86 |
| 146 | G2 | NEC-4 | MPXV | FAM | No Amp | Undetermined |
| 146 | G2 | NEC-4 | RNAse P | VIC | Inconclusive | 38.45 |
| 147 | G3 | MPX-8 | MPXV | FAM | Amp | 34.42 |
| 147 | G3 | MPX-8 | RNAse P | VIC | Amp | 15.85 |
| 148 | G4 | NEC-12 | MPXV | FAM | No Amp | Undetermined |
| 148 | G4 | NEC-12 | RNAse P | VIC | Amp | 37.56 |
| 150 | G6 | NEC-16 | MPXV | FAM | No Amp | Undetermined |
| 150 | G6 | NEC-16 | RNAse P | VIC | Inconclusive | 38.52 |
| 152 | G8 | MPX-20 | MPXV | FAM | Amp | 34.64 |
| 152 | G8 | MPX-20 | RNAse P | VIC | Amp | 15.85 |
| 154 | G10 | NEC-24 | MPXV | FAM | No Amp | Undetermined |
| 154 | G10 | NEC-24 | RNAse P | VIC | No Amp | Undetermined |
| 170 | H2 | MPX-4 | MPXV | FAM | Amp | 34.40 |
| 170 | H2 | MPX-4 | RNAse P | VIC | Amp | 16.19 |
| 171 | H3 | NEC-8 | MPXV | FAM | No Amp | Undetermined |
| 171 | H3 | NEC-8 | RNAse P | VIC | No Amp | Undetermined |
| 172 | H4 | MPX-12 | MPXV | FAM | Amp | 33.89 |
| 172 | H4 | MPX-12 | RNAse P | VIC | Amp | 15.70 |
| 174 | H6 | MPX-16 | MPXV | FAM | Amp | 35.01 |
| 174 | H6 | MPX-16 | RNAse P | VIC | Amp | 15.59 |
| 176 | H8 | NEC-20 | MPXV | FAM | Inconclusive | 19.00 |
| 176 | H8 | NEC-20 | RNAse P | VIC | No Amp | Undetermined |
| 178 | H10 | MPX-24 | MPXV | FAM | Amp | 35.86 |
| 178 | H10 | MPX-24 | RNAse P | VIC | Amp | 15.93 |
| 180 | H12 | PC | MPXV | FAM | Amp | 25.68 |
| 180 | H12 | PC | RNAse P | VIC | Amp | 26.58 |
| 384 | P24 | NTC | MPXV | FAM | No Amp | Undetermined |
| 384 | P24 | NTC | RNAse P | VIC | No Amp | Undetermined |

**Conclusion:** No cross contamination was observed.

#### 8.7. Cross Reactivity (Analytical Specificity)

Cross-reactivity studies were performed to demonstrate that the test does not react with related pathogens, high prevalence disease agents, and normal or pathogenic flora that are reasonably likely to be encountered in a clinical sample. A wet testing with Similar organism(s) will be performed using higher copies/ml.

Orthopox family virus was used to conduct the studies for the Cross reactivity. Cowpox virus, Camel pox virus and rabbit pox virus were used for the studies.

**Results:**

| Sample | Target | Amp Status | Cq |
|---|---|---|---|
| CowpoxVirus-1 | MPXV | Amp | 32.02 |
| CowpoxVirus-1 | RNAse P | Amp | 23.55 |
| CowpoxVirus-2 | MPXV | Amp | 33.12 |
| CowpoxVirus-2 | RNAse P | Amp | 24.08 |
| CowpoxVirus-3 | MPXV | Amp | 32.54 |
| CowpoxVirus-3 | RNAse P | Amp | 23.65 |
| MPXVirus-1 | MPXV | Amp | 27.14 |
| MPXVirus-1 | RNAse P | Amp | 26.35 |
| CamelpoxVirus-1 | MPXV | Amp | 29.90 |
| CamelpoxVirus-1 | RNAse P | Amp | 16.81 |
| CamelpoxVirus-2 | MPXV | Amp | 30.90 |
| CamelpoxVirus-2 | RNAse P | Amp | 16.75 |
| CamelpoxVirus-3 | MPXV | Amp | 30.72 |
| CamelpoxVirus-3 | RNAse P | Amp | 16.69 |
| RabbitpoxVirus-1 | MPXV | No Amp | Undetermined |
| RabbitpoxVirus-1 | RNAse P | Amp | 38.03 |
| RabbitpoxVirus-2 | MPXV | No Amp | Undetermined |
| RabbitpoxVirus-2 | RNAse P | No Amp | Undetermined |
| RabbitpoxVirus-3 | MPXV | No Amp | Undetermined |
| RabbitpoxVirus-3 | RNAse P | No Amp | Undetermined |
| PC | MPXV | Amp | 24.47 |
| PC | RNAse P | Amp | 25.16 |
| NTC-2 | MPXV | No Amp | Undetermined |
| NTC-2 | RNAse P | Amp | 38.01 |
| NTC-1 | MPXV | No Amp | Undetermined |
| NTC-1 | RNAse P | Inconclusive | 37.86 |

**Conclusion:**

The assay can detect related orthopoxviruses, including Monkeypox virus. Further testing will be done to confirm other orthopoxviruses. Rabbitpox will be evaluated further.

**8.8. Interfering substances**

Pooled negative saliva specimens will be spiked with known positive sample at 2x LoD and potential interfering substances at the concentrations below. Each substance will be tested in 10 replicates. Any test substance that fails to meet acceptance criteria may be evaluated at lower concentrations to establish the concentration at which the substance is tolerated.

| Interfering Substance | Concentration |
|---|---|
| Blood (Human) -B | 1% v/v |
| Colgate toothpaste-T | 0.25% v/v |
| Cepacol throat lozenges (menthol + Benzocaine)-C | 5 mg/mL |
| Crest Mouthwash-M | 2.5% v/v |
| Human DNA-DNA | 10ng/uL |

Acceptance criteria for interfering substances:

1. ≥90% of replicates are positive for MPXV specimens. If less than 90% are in agreement, the substance will be diluted to determine the concentration that is tolerated by the assay.

**Results**

| Sample | Target | Amp Status | Cq |
|---|---|---|---|
| NEC-1 | MPXV | No Amp | Undetermined |
| NEC-1 | RNAse P | Amp | 36.79 |
| MPXV_B-1 | MPXV | Amp | 34.02 |
| MPXV_B-1 | RNAse P | Amp | 16.31 |
| MPXV_B-2 | MPXV | Amp | 34.96 |
| MPXV_B-2 | RNAse P | Amp | 15.43 |
| MPXV_B-3 | MPXV | Amp | 33.64 |
| MPXV_B-3 | RNAse P | Amp | 15.71 |
| MPXV_B-4 | MPXV | Amp | 35.92 |
| MPXV_B-4 | RNAse P | Amp | 15.73 |
| MPXV_B-5 | MPXV | Amp | 36.75 |
| MPXV_B-5 | RNAse P | Amp | 15.88 |
| MPXV_B-6 | MPXV | Amp | 36.03 |
| MPXV_B-6 | RNAse P | Amp | 15.85 |
| MPXV_B-7 | MPXV | Amp | 35.45 |
| MPXV_B-7 | RNAse P | Amp | 15.65 |
| MPXV_B-8 | MPXV | Amp | 34.12 |
| MPXV_B-8 | RNAse P | Amp | 16.96 |
| MPXV_B-9 | MPXV | No Amp | Undetermined |
| MPXV_B-9 | RNAse P | Amp | 15.70 |
| MPXV_B-10 | MPXV | Amp | 35.96 |
| MPXV_B-10 | RNAse P | Amp | 16.02 |
| NEC-2 | MPXV | No Amp | Undetermined |
| NEC-2 | RNAse P | No Amp | Undetermined |

| | | | |
|---|---|---|---|
| MPXV-T-1 | MPXV | Amp | 35.47 |
| MPXV-T-1 | RNAse P | Amp | 16.85 |
| MPXV-T-2 | MPXV | Amp | 36.08 |
| MPXV-T-2 | RNAse P | Amp | 16.48 |
| MPXV-T-3 | MPXV | Amp | 36.93 |
| MPXV-T-3 | RNAse P | Amp | 16.77 |
| MPXV-T-4 | MPXV | Amp | 36.79 |
| MPXV-T-4 | RNAse P | Amp | 16.95 |
| MPXV-T-5 | MPXV | Amp | 36.02 |
| MPXV-T-5 | RNAse P | Amp | 16.13 |
| MPXV-T-6 | MPXV | Amp | 32.77 |
| MPXV-T-6 | RNAse P | Amp | 15.91 |
| MPXV-T-7 | MPXV | Amp | 33.70 |
| MPXV-T-7 | RNAse P | Amp | 15.57 |
| MPXV-T-8 | MPXV | Amp | 32.44 |
| MPXV-T-8 | RNAse P | Amp | 15.26 |
| MPXV-T-9 | MPXV | Amp | 22.50 |
| MPXV-T-9 | RNAse P | Amp | 15.23 |
| MPXV-T-10 | MPXV | Amp | 21.20 |
| MPXV-T-10 | RNAse P | Amp | 15.01 |
| NEC-3 | MPXV | No Amp | Undetermined |
| NEC-3 | RNAse P | Amp | 38.11 |
| MPXV-M-1 | MPXV | Amp | 35.59 |
| MPXV-M-1 | RNAse P | Amp | 16.35 |
| MPXV-M-2 | MPXV | Amp | 32.90 |
| MPXV-M-2 | RNAse P | Amp | 16.22 |
| MPXV-M-3 | MPXV | Amp | 35.72 |
| MPXV-M-3 | RNAse P | Amp | 16.21 |
| MPXV-M-4 | MPXV | Amp | 34.62 |
| MPXV-M-4 | RNAse P | Amp | 16.16 |
| MPXV-M-5 | MPXV | Amp | 34.15 |
| MPXV-M-5 | RNAse P | Amp | 16.15 |
| MPXV-M-6 | MPXV | Amp | 34.11 |
| MPXV-M-6 | RNAse P | Amp | 16.04 |
| MPXV-M-7 | MPXV | Amp | 33.54 |
| MPXV-M-7 | RNAse P | Amp | 16.14 |
| MPXV-M-8 | MPXV | Amp | 33.87 |
| MPXV-M-8 | RNAse P | Amp | 16.17 |
| MPXV-M-9 | MPXV | Amp | 34.58 |
| MPXV-M-9 | RNAse P | Amp | 16.10 |
| MPXV-M-10 | MPXV | Amp | 33.86 |
| MPXV-M-10 | RNAse P | Amp | 16.07 |
| MPXV_C-1 | MPXV | Amp | 35.65 |
| MPXV_C-1 | RNAse P | Amp | 15.87 |
| MPXV_C-2 | MPXV | Amp | 37.13 |
| MPXV_C-2 | RNAse P | Amp | 16.18 |
| MPXV_C-3 | MPXV | Amp | 34.32 |
| MPXV_C-3 | RNAse P | Amp | 15.74 |
| MPXV_C-4 | MPXV | Amp | 35.60 |
| MPXV_C-4 | RNAse P | Amp | 15.66 |
| MPXV_C-5 | MPXV | Amp | 35.97 |
| MPXV_C-5 | RNAse P | Amp | 15.83 |
| MPXV_C-6 | MPXV | Amp | 36.60 |
| MPXV_C-6 | RNAse P | Amp | 15.90 |

| | | | |
|---|---|---|---|
| MPXV_C-7 | MPXV | Amp | 34.51 |
| MPXV_C-7 | RNAse P | Amp | 16.03 |
| MPXV_C-8 | MPXV | Amp | 36.71 |
| MPXV_C-8 | RNAse P | Amp | 16.20 |
| MPXV_C-9 | MPXV | Amp | 31.81 |
| MPXV_C-9 | RNAse P | Amp | 14.20 |
| MPXV_C-10 | MPXV | No Amp | Undetermined |
| MPXV_C-10 | RNAse P | Amp | 16.28 |
| MPXV_DNA-1 | MPXV | Amp | 35.64 |
| MPXV_DNA-1 | RNAse P | Amp | 16.43 |
| MPXV_DNA-2 | MPXV | Amp | 35.18 |
| MPXV_DNA-2 | RNAse P | Amp | 16.26 |
| MPXV_DNA-3 | MPXV | Amp | 34.79 |
| MPXV_DNA-3 | RNAse P | Amp | 16.25 |
| MPXV_DNA-4 | MPXV | Amp | 35.02 |
| MPXV_DNA-4 | RNAse P | Amp | 16.17 |
| MPXV_DNA-5 | MPXV | Amp | 34.61 |
| MPXV_DNA-5 | RNAse P | Amp | 16.13 |
| MPXV_DNA-6 | MPXV | Amp | 37.58 |
| MPXV_DNA-6 | RNAse P | Amp | 16.30 |
| MPXV_DNA-7 | MPXV | Amp | 35.62 |
| MPXV_DNA-7 | RNAse P | Amp | 16.09 |
| MPXV_DNA-8 | MPXV | Amp | 35.19 |
| MPXV_DNA-8 | RNAse P | Amp | 16.18 |
| MPXV_DNA-9 | MPXV | Amp | 35.51 |
| MPXV_DNA-9 | RNAse P | Amp | 16.12 |
| MPXV_DNA-10 | MPXV | No Amp | Undetermined |
| MPXV_DNA-10 | RNAse P | Amp | 16.79 |
| NTC | MPXV | No Amp | Undetermined |
| NTC | RNAse P | No Amp | Undetermined |
| PC-1 | MPXV | Amp | 18.88 |
| PC-1 | RNAse P | Amp | 19.93 |

**Conclusion:**

Each substance was tested at the given standard concentrations. No interference effects were observed for any of the substances at the concentrations tested except for blood and human DNA. One sample spiked with blood and another with human DNA did not show amplification. However, the study was still within the 95% acceptable range so no further testing was required.

**8.9. Stability**

The known positive samples were prepared at two different dilutions and stored at room temperature and 4 degree celcius over the course of 8 days (0, 1, 4, 8 days). Data for day 0 and day 1 is shown below. The report will be updated when data for the remainder of the days is available.

| | | RT | | | | RT | |
|---|---|---|---|---|---|---|---|
| | | DAY 0 | | | | DAY 1 | |
| Sample | Target | Amp Status | Cq | | Sample | Amp Status | Cq |
| NEC-33 | MPXV | No Amp | Undetermined | | NEC1 | No Amp | Undetermined |
| NEC-33 | RNAse P | Inconclusive | 38.53 | | NEC1 | No Amp | Undetermined |
| NEC-34 | MPXV | No Amp | Undetermined | | NEC2 | No Amp | Undetermined |
| NEC-34 | RNAse P | No Amp | Undetermined | | NEC2 | Inconclusive | 37.52 |

| | | | | | | | |
|---|---|---|---|---|---|---|---|
| NEC-35 | MPXV | No Amp | Undetermined | | NEC3 | Inconclusive | 39.84 |
| NEC-35 | RNAse P | Inconclusive | 38.25 | | NEC3 | Inconclusive | 38.41 |
| Patient2-1x-0-1 | MPXV | Amp | 23.56 | | Patient2-1x-RT-1 | Amp | 23.66 |
| Patient2-1x-0-1 | RNAse P | Amp | 15.13 | | Patient2-1x-RT-1 | Amp | 16.42 |
| Patient2-1x-0-2 | MPXV | Amp | 23.53 | | Patient2-1x-RT-2 | Amp | 23.93 |
| Patient2-1x-0-2 | RNAse P | Amp | 15.17 | | Patient2-1x-RT-2 | Amp | 16.33 |
| Patient2-1x-0-3 | MPXV | Amp | 23.45 | | Patient2-1x-RT-3 | Amp | 24.20 |
| Patient2-1x-0-3 | RNAse P | Amp | 15.17 | | Patient2-1x-RT-3 | Amp | 16.34 |
| Patient2-2x-0-1 | MPXV | Amp | 24.71 | | Patient2-2x-RT-1 | Amp | 25.09 |
| Patient2-2x-0-1 | RNAse P | Amp | 15.00 | | Patient2-2x-RT-1 | Amp | 16.00 |
| Patient2-2x-0-2 | MPXV | Amp | 24.33 | | Patient2-2x-RT-2 | Amp | 24.90 |
| Patient2-2x-0-2 | RNAse P | Amp | 14.86 | | Patient2-2x-RT-2 | Amp | 15.76 |
| Patient2-2x-0-3 | MPXV | Amp | 24.53 | | Patient2-2x-RT-3 | Amp | 25.09 |
| Patient2-2x-0-3 | RNAse P | Amp | 15.05 | | Patient2-2x-RT-3 | Amp | 16.02 |
| Negative Saliva-0-1 | MPXV | No Amp | Undetermined | | NegSaliva-1-1 | No Amp | Undetermined |
| Negative Saliva-0-1 | RNAse P | Amp | 14.92 | | NegSaliva-1-1 | Amp | 16.33 |
| Negative Saliva-0-2 | MPXV | No Amp | Undetermined | | NegSaliva-1-2 | No Amp | Undetermined |
| Negative Saliva-0-2 | RNAse P | Amp | 14.99 | | NegSaliva-1-2 | Amp | 16.23 |
| Negative Saliva-0-3 | MPXV | No Amp | Undetermined | | NegSaliva-1-3 | No Amp | Undetermined |
| Negative Saliva-0-3 | RNAse P | Amp | 14.80 | | NegSaliva-1-3 | Amp | 16.22 |
| Negative Saliva -0-4 | MPXV | Inconclusive | 24.95 | | NegSaliva-1--4 | No Amp | Undetermined |
| Negative Saliva -0-4 | RNAse P | Amp | 14.84 | | NegSaliva-1--4 | Amp | 15.94 |
| | | **4 °C** | | | | **4 °C** | |
| | | **DAY 0** | | | | **DAY 1** | |
| **Sample** | **Target** | **Amp Status** | **Cq** | | **Sample** | **Amp Status** | **Cq** |
| NEC-36 | MPXV | Inconclusive | 21.37 | | NEC4 | No Amp | Undetermined |
| NEC-36 | RNAse P | Inconclusive | 15.48 | | NEC4 | No Amp | Undetermined |
| NEC-37 | MPXV | No Amp | Undetermined | | NEC5 | No Amp | Undetermined |
| NEC-37 | RNAse P | No Amp | Undetermined | | NEC5 | No Amp | Undetermined |
| NEC-38 | MPXV | Inconclusive | 37.05 | | NEC6 | No Amp | Undetermined |
| NEC-38 | RNAse P | Inconclusive | 27.88 | | NEC6 | Amp | 36.77 |
| Patient2-1x-0-1-4C | MPXV | Amp | 23.65 | | Patient2-1x-4C-1 | Amp | 23.90 |
| Patient2-1x-0-1-4C | RNAse P | Amp | 15.52 | | Patient2-1x-4C-1 | Amp | 16.06 |
| Patient2-1x-0-2-4C | MPXV | Amp | 22.81 | | Patient2-1x-4C-2 | Amp | 23.79 |
| Patient2-1x-0-2-4C | RNAse P | Amp | 15.17 | | Patient2-1x-4C-2 | Amp | 15.98 |
| Patient2-1x-0-3-4C | MPXV | Amp | 23.88 | | Patient2-1x-4C-3 | Amp | 23.94 |
| Patient2-1x-0-3-4C | RNAse P | Amp | 15.25 | | Patient2-1x-4C-3 | Amp | 16.16 |
| Pateint2-2x-0-1-4C | MPXV | Amp | 24.61 | | Patient2-2x-4C-1 | Amp | 24.88 |
| Pateint2-2x-0-1-4C | RNAse P | Amp | 15.10 | | Patient2-2x-4C-1 | Amp | 15.96 |
| Pateint2-2x-0-2-4C | MPXV | Amp | 24.74 | | Patient2-2x-4C-2 | Amp | 24.76 |
| Pateint2-2x-0-2-4C | RNAse P | Amp | 15.00 | | Patient2-2x-4C-2 | Amp | 15.94 |
| Pateint2-2x-0-3-4C | MPXV | Amp | 24.28 | | Patient2-2x-4C-3 | Amp | 24.53 |
| Pateint2-2x-0-3-4C | RNAse P | Amp | 14.89 | | Patient2-2x-4C-3 | Amp | 16.06 |
| Negative Saliva-1-0 | MPXV | No Amp | Undetermined | | NegSaliva-1-01 | No Amp | Undetermined |
| Negative Saliva-1-0 | RNAse P | Amp | 13.98 | | NegSaliva-1-01 | Amp | 14.68 |
| Negative Saliva-2-0 | MPXV | No Amp | Undetermined | | NegSaliva-01-2 | No Amp | Undetermined |
| Negative Saliva-2-0 | RNAse P | Amp | 14.14 | | NegSaliva-01-2 | Amp | 14.98 |
| Negative Saliva-3-0 | MPXV | No Amp | Undetermined | | NegSaliva-01-3 | No Amp | Undetermined |
| Negative Saliva-3-0 | RNAse P | Amp | 14.06 | | NegSaliva-01-3 | Amp | 14.99 |
| Negative Saliva-4-0 | MPXV | No Amp | Undetermined | | NegSaliva-01-4 | No Amp | Undetermined |
| Negative Saliva-4-0 | RNAse P | Amp | 14.11 | | NegSaliva-01-4 | Amp | 14.98 |
| PC-1 | MPXV | Amp | 18.88 | | PC-1 | Amp | 25.58 |
| PC-1 | RNAse P | Amp | 19.93 | | PC-1 | Amp | 26.60 |
| NTC1 | MPXV | No Amp | Undetermined | | NTC2 | No Amp | Undetermined |
| NTC1 | RNAse P | Inconclusive | 37.85 | | NTC2 | Inconclusive | 38.09 |

**Conclusion:**

No significance difference was observed on day0 and day1-Sample was stable.
